# Spatially informed comprehensive tumor transcriptomic profiling stratifies clinical outcomes in early triple negative breast cancer

**DOI:** 10.64898/2026.07.06.26357224

**Authors:** Barbora Huraiová, Michal Gala, Liliane Barroso, Anna Lea Amylidi, Daniela Gábrišová, Soňa Gubová, Tomáš Ondris, Kierann Javorčík, Marek Kucej, Filip Németh, Maximilián Rada, Simona Smolková, Ema Husarčíková, Natália Matyašovská, Adrián Szobi, Sandra Szeibeczederová, Alexandra Čapkovičová, Zoltán Ferjentsik, Sára Hrabovská, Iveta Vereš, Hande Özbaşak, Steven A. Calle, Peter Grell, Miloš Holánek, Rudolf Nenutil, Iveta Selingerová, François Cherifi, George Emile, Roman Rouzier, Peter Regitnig, Karl Tamussino, Katarzyna J. Jerzak, Fang-I Lu, Sunitha Shetty, Laura Comerma, Joan Albanell, Sonia Servitja, Igor Andrašina, David A. Eberhard, Konstantinos Papazisis, Gabriel Rinnerthaler, Evan D. Paul, Pavol Čekan

## Abstract

The intensification of neoadjuvant therapy for early triple-negative breast cancer (eTNBC) – through the addition of carboplatin to standard chemotherapy and the incorporation of pembrolizumab – has markedly improved prognosis in recent years. However, this escalation carries a substantial risk of toxicity, and not all patients require the full regimen to achieve benefit. Realizing individualized treatment strategies will therefore depend on prognostic and predictive biomarkers that can forecast treatment response and long-term outcome. In the present study, we interrogated public gene expression datasets to develop transcriptomic signatures predicting response to neoadjuvant treatment and risk of recurrence. To validate these signatures, we used the Multiplex8+ platform for spatially informed comprehensive transcriptomic profiling in a real-world, multicenter, retrospective cohort of 590 patients diagnosed with eTNBC and treated with neoadjuvant chemotherapy with or without immunotherapy. The diagnostic Multiplex8+ test uses H&E and multiplexed RNA-FISH to guide the selection of specific tumor areas for the whole transcriptome sequencing and signature analysis. In the real-world cohort, the Multiplex8+ signatures were associated with both response and prognosis, remaining highly significant in multivariable models that included clinical parameters. The signatures were complementary to established biomarkers such as stromal tumor-infiltrating lymphocytes. These findings warrant prospective integration of the signatures into risk-stratified clinical trials to support future de-escalation and escalation strategies, enabling a better balance of efficacy, toxicity, cost, and drug availability.

## Introduction

Triple negative breast cancers (TNBC) comprise 10-20% of all breast cancers, and tend to be biologically aggressive, frequently high grade, and are associated with worse prognosis. TNBC disproportionately afflicts young women, minorities, and women carrying *BRCA1* mutations^1–3^. Efforts to increase early breast cancer screening in women at risk are expected to increase the frequency of diagnosis of early-stage TNBC (eTNBC), holding promise of improved long-term outcomes through earlier detection and enabling curative intent therapy. Typically, patients with high-risk eTNBC receive neoadjuvant chemoimmunotherapy, followed by surgery and additional systemic therapy post-operatively, which is guided by the presence or absence of residual disease. Pathological tumor response assessed in the surgical specimen and regional lymph nodes allows for prognostic stratification and guidance of risk adapted adjuvant therapy. Patients achieving a complete pathological response (pCR) generally have an excellent prognosis with very low recurrence rates^4^.

Options for systemic therapy for TNBC are evolving. TNBC is defined by the lack of expression of estrogen and progesterone hormone receptors (HR) and lack of overexpression and amplification of the HER2 protein and its oncogene (*ERBB2*) oncogene, respectively. TNBC is a heterogeneous group of breast cancers, and a unifying molecular driver has not been defined. Thus, the mainstay of TNBC treatment has been various cytotoxic chemotherapy (CTx) regimens^5,6^. The relatively recent addition of immunotherapy based on the pivotal KEYNOTE-522 (KN-522) trial, which evaluated adding neoadjuvant pembrolizumab to a taxane-carboplatin-anthracycline/cyclophosphamide backbone followed by adjuvant pembrolizumab, demonstrated a pCR improvement from 51.2% to 64.8%^7^ as well as prolonged event-free (EFS)^8^ and overall survival (OS)^9^. Based on these results, the KN-522 regimen has become standard of care (SoC) for stage II-III eTNBC^10–13^.

Immune checkpoint inhibitors and cytotoxic chemotherapies provide meaningful clinical benefit in eTNBC, but also present significant clinical and financial toxicities. Post-approval real-world studies confirm that the KN-522 regimen improves pCR rates but also report high rates of severe adverse events, drug dose reductions or discontinuation, and medical contacts^14–20^.

Neoadjuvant decision-making in eTNBC would benefit substantially from tests that predict the likelihood of benefit from ICIs or specific CTx regimens, enabling treatment to be tailored to the individual patient — escalating therapy for those unlikely to respond to less intensive regimens, while sparing patients with low-risk the toxicity of unnecessary treatment. In unresectable or metastatic TNBC, clinical benefit of pembrolizumab is seen in those patients whose tumors are PD-L1 positive by IHC^21,22^, defining the approved indication for use. However, PD-L1 IHC was not predictive of eTNBC response or survival benefit in KN-522^7,8^. Recently, stromal tumor-infiltrating lymphocytes (TILs) have been explored as possible biomarkers of response to ICI in eTNBC^23–25^ but the predictive specificity may be poor, since TILs are associated with better responses and prognosis irrespective of specific therapy^26,27^.

Gene expression profiling is established as a powerful approach that has been clinically validated and adopted for diagnostic classification, prognostication and treatment prediction in breast cancer^28–30^. An important limitation to the clinical accuracy of these molecular assays is the heterogeneous cellular composition of tumors, including tumor cells and a variety of non-tumor cell types – stroma, immune, vascular – and even within the tumor cell population itself. Various “spatial biology” and single cell platforms have been developed to study tumor heterogeneity in research settings. In clinical diagnostics, solid tumor heterogeneity is addressed by establishing a minimum tumor cell content needed for a successful assay, and by enriching the tumor cell content using a dissection technique. Manual macrodissection is simpler but less precise than microdissection using a microscope to visualize and capture tissue regions at the cellular level of discrimination.

Multiplex8+ is a spatially-informed gene expression profiling platform that maximizes accuracy and versatility for solid tumor testing^31^. Serial adjacent FFPE tissue sections provide H&E staining and multiplex in situ fluorescence imaging of selected gene transcript panels (mRNA-FISH). Digital and molecular histology are used to inform laser-capture microdissection (LCM) of desired cell areas, from which mRNA is isolated for whole transcriptome RNAseq. Multiplex8+ permits gene expression profiling of tumor cells enriched from small heterogenous samples, such as core needle biopsies, and is a versatile platform for developing and testing gene signatures related to clinical prognosis, prediction, diagnosis, tumor-immune biology. Next to CTx response prediction, this can be especially useful to predict benefit from therapeutics such as antibody drug conjugates (ADCs) and ICIs, where expression of a single biomarker is insufficient to address their complex mechanisms of action.

In the present study, we developed predictive and prognostic signatures for patients with eTNBC treated with chemotherapy with or without immunotherapy and validated their performance using Multiplex8+ in a large real-world cohort.

## Results

### Development of Multiplex8+ signatures to stratify clinical outcomes in patients with eTNBC

We mined publicly available gene expression datasets to develop three distinct gene signature classifiers in eTNBC: (1) a predictive signature for sensitivity to neoadjuvant chemotherapy (MDX-CTx), (2) a predictive signature for response to immunotherapy (MDX-ICI), and (3) a prognostic signature for recurrence risk (MDX-Risk). A detailed overview of our approach and results is provided in the **Supplementary information** and briefly described here (**Fig. 1a**). To identify gene signatures that predict sensitivity to neoadjuvant treatment, we used a modified machine learning (ML) pipeline incorporating stratified 5-fold cross-validation, supervised feature (gene) selection, and an iterative model fitting strategy^32^. We applied this pipeline on the Sotiriou dataset^33^, a curated compendium of microarray data from 23 studies involving 706 patients with eTNBC who received neoadjuvant therapy, to identify a 45-gene logistic regression gene expression signature related to chemotherapy sensitivity (MDX-CTx). To develop an immunotherapy response signature, we used the pembrolizumab (n=29) and the durvalumab plus olaparib (n=21) arms of I-SPY2^34,35^ and our feature space consisted of a curated list of genes associated with neoadjuvant treatment response and survival outcomes obtained from a large TNBC transcriptome meta-analysis^36^. We used stratified 3-fold cross-validation and LASSO logistic regression to define a 20-gene logistic regression gene expression signature related to chemoimmunotherapy sensitivity (MDX-ICI).

**Figure 1.**
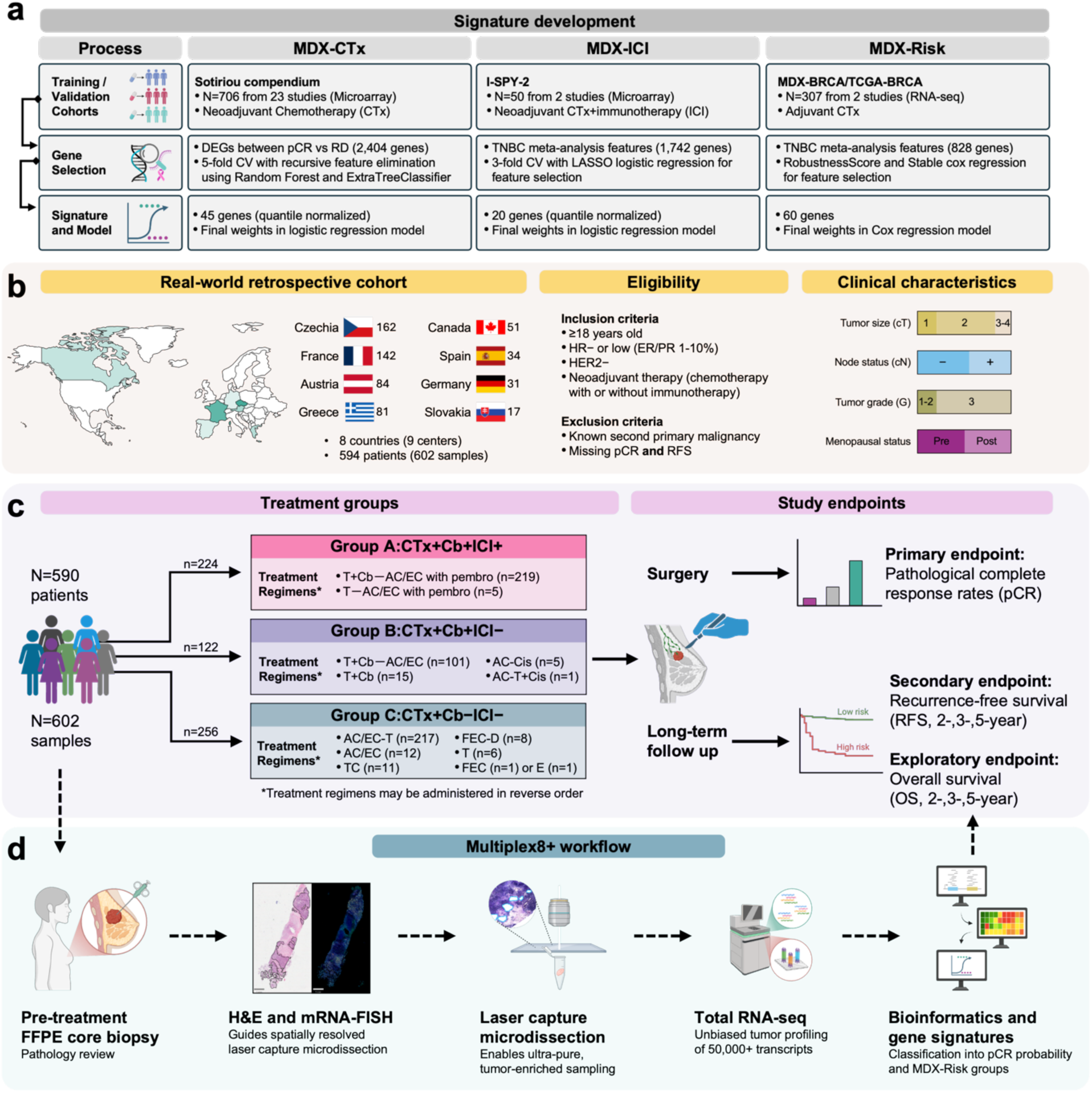
The schematic provides an overview on the development and validation of gene/signatures, the study sites and eligibility criteria, study design, and Multiplex8+ workflow. **a**) The three gene signatures used in this study were developed and validated on 1,063 TNBC patients from publicly available datasets using a 3-step process: 1) identification, retrieval, and processing of relevant datasets; 2) curation of top genes followed by cross validation and gene feature selection; and 3) model fitting and derivation of gene sets, algorithms, and weights for final signatures. **b**) The map shows the countries of the nine cancer centers that provided core biopsies and linked clinical data included in the real-world study. The flags depict each country and the number of patients. The eligibility section lists the inclusion and exclusion criteria for the study. The clinical characteristics section highlights key baseline clinical parameters for the overall cohort. **c**) Study design illustrates the number of patients and samples, stratification into three treatment groups, the composition of treatment regimens in each group, and primary, secondary, and exploratory clinical endpoints. **d**) Diagnostic FFPE core biopsies from all eligible patients were process using the Multiplex8+ platform, which consists of pathology review, H&E and multiplexed RNA-FISH to identify and annotate spatial characteristics, laser capture microdissection of spatially defined tumor regions, unbiased whole transcriptome sequencing, followed by bioinformatics, calculation of signature scores, and construction of higher order groups to associate with clinical endpoints. AC/EC, anthracyclines (Adriamycin or Epirubicin) and cyclophosphamide; CTx, chemotherapy regimen, ICI, immunotherapy regimen; Cb, carboplatin; Cis, cisplatin; CV, cross-validation; DEGs, differentially expressed genes; ER, estrogen receptor; F, 5-Fluorouracil; H&E, hematoxylin & eosin; HR, hormone receptors; mRNA-FISH, multiplexed RNA-fluorescent in situ hybridization; PR, progesterone receptor; T, taxanes (paclitaxel or docetaxel). Some images and icons were created in BioRender. Paul, E. (2026) https://BioRender.com/xw5i8gj. Flag icons were obtained from Wikimedia Commons in the public domain or under a free-to-use CC0 license. The world map and borders were obtained from the public domain Natural Earth.

To develop and validate a prognostic signature we started with a curated list of genes from the TNBC transcriptome meta-analysis^36^. Then, we used stable Cox regression^37^ to distill these features into a 60-gene Cox regression model (MDX-Risk) using two large international cohorts of patients with eTNBC treated in the adjuvant setting: MDX-BRCA^31^ (training set, n=159) and TCGA-BRCA^38,39^ (validation set, n=148).

### Signature validation in a real-world, multicenter, retrospective cohort of eTNBC

To validate the predictive and prognostic abilities of our gene signatures, we retrieved 657 diagnostic pre-treatment core biopsies and associated clinicopathological data from 634 patients with eTNBC treated across seven European countries and Canada (**Fig. 1b**). The patients were treated in the neoadjuvant setting between April 2006 to February 2026 with either (1) chemotherapy (taxane plus carboplatin followed or preceded by anthracycline/cyclophosphamide) combined with the ICI, pembrolizumab (e.g., the KN-522 regimen; group A, CTx+Cb+ICI+), (2) the KN-522 chemotherapy backbone without pembrolizumab (group B, CTx+Cb+ICI−), or (3) chemotherapy (taxane or anthracycline/cyclophosphamide or both without carboplatin and pembrolizumab, group C, CTx+Cb−ICI−). The therapeutic regimens that comprise each group and the number of patients who received a specific regimen are listed in the **Methods** (also see **Fig. 1c**). As shown in the CONSORT diagram (**Extended Data Fig. 1)**, after excluding 50 samples for pre-analytical reasons (i.e., ineligible, revoked informed consent, no tumor), we processed the remaining 608 samples (594 patients) with our spatially-informed Multiplex8+ workflow (**Fig. 1d**)^31^. Following exclusion of technical dropouts, this yielded a final cohort of 602 samples (590 patients) for correlative analyses of gene signatures with pCR (primary endpoint) and 2-,3-, or 5-year recurrence-free survival (RFS, secondary endpoint) and overall survival (OS, exploratory endpoint) depending on the group.

Baseline characteristics of the patients in each of the three treatment groups and the overall cohort are shown in **Table 1**. Overall, the median age was 50.5 (23-86-year range), 80% had tumors larger than 2 cm (cT2-4), 55% were node negative, 75% had a tumor grade 3, 30% had HER2 low (IHC 1+/2+ with equivocal results confirmed with DNA-FISH/SISH/ISH as non-amplified) status, and 8% were classified as HR-low positive (estrogen and/or progesterone receptor expression between 1-10%). Clinical parameters were similar between treatment groups with most differences ≤5%; however, the CTx+Cb+ICI− group contained more cT1 and fewer cT2 tumors and the CTx+Cb+ICI+ group had more G2 tumors. Expectedly, given the historical differences in treatment regimens, the CTx+Cb−ICI− group had the longest median follow-up time (57 months) followed by the CTx+Cb+ICI− group (44 months), and then the CTx+Cb+ICI+ group (19 months). Data missingness was minimal (≤8%) for most clinical parameters and evenly distributed among treatment groups.

**Table 1.** Baseline clinical characteristics of the patients.

| Category | CTx+Cb-ICI- | CTx+Cb+ICI- | CTx+Cb+ICI+ | Total |
| --- | --- | --- | --- | --- |
| **Total #** | 256 | 122 | 224 | 602 |
| **Origin** |  |  |  |  |
| *Spain* | 4 (2%) | 16 (13%) | 14 (6%) | 34 (6%) |
| *Czech Replublic* | 75 (29%) | 29 (24%) | 58 (26%) | 162 (27%) |
| *Canada* | 28 (11%) | 1 (1%) | 22 (10%) | 51 (8%) |
| *France* | 66 (26%) | 15 (12%) | 61 (27%) | 142 (24%) |
| *Biobank Graz (AT)* | 58 (23%) | 21 (17%) | 5 (2%) | 84 (14%) |
| *Greece* | 10 (4%) | 24 (20%) | 47 (21%) | 81 (13%) |
| *Slovakia* | 3 (1%) | 4 (3%) | 10 (4%) | 17 (3%) |
| *Leipzig biobank (DE)* | 1 (>1%) | 5 (4%) | 0 (0%) | 6 (1%) |
| *PATH biobank (DE)* | 11 (4%) | 7 (6%) | 7 (3%) | 25 (4%) |
| **Median age in years (range)** | 50.5 (24 – 86) | 50 (23 – 84) | 50.5 (26 – 82) | 50.5 (23 – 86) |
| **Age category** |  |  |  |  |
| *≤40 years* | 53 (21%) | 42 (34%) | 52 (23%) | 147 (24%) |
| *41–49 years* | 58 (23%) | 28 (23%) | 62 (28%) | 148 (25%) |
| *50–64 years* | 73 (29%) | 29 (24%) | 68 (30%) | 170 (28%) |
| *≥65 years* | 72 (28%) | 23 (19%) | 42 (19%) | 137 (23%) |
| **Tumor size** |  |  |  |  |
| *cT1* | 57 (22%) | 29 (24%) | 36 (16%) | 122 (20%) |
| *cT2* | 164 (64%) | 65 (53%) | 145 (65%) | 374 (62%) |
| *cT3* | 22 (9%) | 18 (15%) | 27 (12%) | 67 (11%) |
| *cT4* | 11 (4%) | 9 (7%) | 15 (7%) | 35 (6%) |
| *Missing* | 2 (1%) | 1 (1%) | 1 (>1%) | 4 (1%) |
| **Node status** |  |  |  |  |
| *cN0* | 142 (55%) | 64 (52%) | 123 (55%) | 329 (55%) |
| *cN1* | 102 (40%) | 48 (39%) | 82 (37%) | 232 (39%) |
| *cN2* | 7 (3%) | 5 (4%) | 9 (4%) | 21 (3%) |
| *cN3* | 3 (1%) | 3 (2%) | 8 (4%) | 14 (2%) |
| *Missing* | 2 (1%) | 2 (2%) | 2 (1%) | 6 (1%) |
| **Tumor grade** |  |  |  |  |
| *G1* | 2 (1%) | 0 (0%) | 3 (1%) | 5 (1%) |
| *G2* | 43 (17%) | 15 (12%) | 53 (24%) | 111 (18%) |
| *G3* | 195 (76%) | 99 (81%) | 157 (70%) | 451 (75%) |
| *Missing* | 16 (6%) | 8 (7%) | 11 (5%) | 35 (6%) |
| **Menopausal status** |  |  |  |  |
| *Pre* | 112 (44%) | 67 (55%) | 114 (51%) | 293 (49%) |
| *Peri* | 2 (1%) | 1 (1%) | 1 (>1%) | 4 (1%) |
| *Post* | 138 (54%) | 49 (40%) | 101 (45%) | 288 (48%) |
| *Missing* | 4 (2%) | 5 (4%) | 8 (4%) | 17 (3%) |
| **Histological subtype** |  |  |  |  |
| *IDC* | 244 (94%) | 115 (94%) | 209 (93%) | 568 (94%) |
| *ILC* | 5 (2%) | 0 (0%) | 2 (1%) | 7 (1%) |
| *ACC* | 0 (0%) | 0 (0%) | 1 (>1%) | 1 (>1%) |
| *Apocrine* | 1 (>1%) | 0 (0%) | 1 (>1%) | 2 (>1%) |
| *Medullary* | 1 (>1%) | 0 (0%) | 0 (>1%) | 1 (>1%) |
| *Metaplastic* | 3 (1%) | 2 (2%) | 1 (>1%) | 6 (1%) |
| *Other* | 2 (1%) | 2 (2%) | 3 (1%) | 7 (1%) |
| *Missing* | 0 (0%) | 2 (2%) | 7 (3%) | 9 (1%) |
| **PR positive (low)** | 3 (1%) | 1 (1%) | 9 (4%) | 13 (2%) |
| **ER positive (low)** | 13 (5%) | 4 (3%) | 15 (7%) | 32 (5%) |
| **HER2 status** |  |  |  |  |
| 2+ | 26 (10%) | 4 (3%) | 22 (10%) | 52 (9%) |
| 1+ | 49 (19%) | 29 (24%) | 48 (21%) | 126 (21%) |
| 0 (or negative) | 179 (70%) | 87 (71%) | 154 (69%) | 420 (70%) |
| *Missing* | 2 (1%) | 2 (2%) | 0 (0%) | 4 (1%) |
| **Ki67 %**<br>**of positive cells**<br>**(range)** | 70%<br>(5% – 99%) | 70%<br>(12% –100%) | 61%<br>(10% –100%) | 70%<br>(5% –100%) |
| **Recurrence event** |  |  |  |  |
| Yes | 63 (25%) | 22 (18%) | 30 (13%) | 115 (19%) |
| No | 170 (66%) | 92 (75%) | 179 (80%) | 441 (73%) |
| *Missing* | 23 (9%) | 8 (7%) | 15 (7%) | 46 (8%) |
| **Median RFS months**<br>**(range)** | 29 (2 – 204) | 30 (1 – 128) | 30 (1 – 87) | 29 (1 – 204) |
| **Deceased** |  |  |  |  |
| Yes | 56 (22%) | 13 (11%) | 13 (6%) | 82 (14%) |
| No | 181 (71%) | 105 (86%) | 192 (86%) | 478 (79%) |
| *Missing* | 19 (7%) | 4 (3%) | 19 (8%) | 42 (7%) |
| **Median OS months**<br>**(range)** | 37 (7 – 204) | 37 (6 – 134) | 37 (1 – 87) | 37 (1 – 204) |
| **Median follow up**<br>**time months (range)** | 55 (6 – 204) | 46 (6 – 134) | 19 (1 – 87) | 36 (1 – 204) |
| **Median year of start**<br>**of neoadjuvant**<br>**treatment (range)** | 2021<br>(2006 – 2024) | 2021<br>(2014 – 2025) | 2021<br>(2018 – 2026) | 2021<br>(2006 – 2026) |

### MDX pCR probability signatures are associated with pathological complete response

Both pCR data and transcriptomic data were available for 600 patients, enabling a correlative analysis of the primary endpoint pCR with the three Multiplex8+ signatures, which stratified patients into biomarker positive and negative subgroups. The pCR rates in the three treatment groups were 59.5% (132/222: CTx+Cb+ICI+ group), 50.8% (62/122: CTx+Cb+ICI− group), and 37.1% (95/256: CTx+Cb−ICI−group), with an overall pCR rate of 48.2% (289/600). In all treatment groups, the biomarker positive subgroups were associated with higher pCR rates (**Extended Data Fig. 2**). The MDX-ICI signature performed strongest in the CTx+Cb+ICI+ group (Δ32.4% difference between MDX-ICI+ and MDX-ICI−) compared to the CTx+Cb+ICI− and CTx+Cb−ICI− groups (Δ28.2% and Δ23.9% differences between MDX-CTx+ and MDX-CTx−); similarly, the MDX-CTx signature showed the largest separation in the CTx+Cb−ICI− and CTx+Cb+ICI− groups (Δ36.3% and Δ31.4%), relative to the CTx+Cb+ICI+ group (Δ22.8%). Although these results are consistent with the signatures performing best in the context of the treatments used in their training, for the most part, the predictive power of these signatures was not treatment specific.

Patients positive for both, the MDX-ICI and the MDX-CTx signature, had the highest pCR rates in all groups, while patients negative for both MDX-ICI and MDX-CTx signature had the lowest pCR rates (**Extended Data Fig. 3**). Patients positive in one signature but negative in the other showed intermediate pCR rates with minimal differences between the MDX-ICI+/CTx− and MDX-ICI−/CTx+ groups. Further combining the two intermediate subgroups resulted in three MDX pCR probability groups: MDX high pCR (MDX-ICI+/CTx+), MDX intermediate pCR (MDX-ICI+/CTx–and MDX-ICI−/CTx+), and a MDX low pCR (MDX-ICI−/CTx−) subgroup.

These three MDX pCR probability groups show marked differences in pCR rates. The MDX pCR high group had increased pCR rates compared to the MDX pCR low group in all three treatment groups: group A (80% vs 30%, OR: 9.01; 95% CI: 4.03-19.37, *p* < 0.0001), group B (84% vs 27%, OR: 14.85; 95% CI: 4.36-49.27, *p* < 0.0001), and group C (69% vs 16%, OR: 11.45; 95% CI: 5.04-24.16, *p* < 0.0001; **Fig. 2a**). A univariable logistic regression showed significantly different odds ratios (OR) for pCR in both the MDX intermediate pCR (OR: 2.70; 95% CI: 1.64-4.62, *p* < 0.0001) and MDX high pCR (OR: 11.54; 95% CI: 6.76-20.38, *p* < 0.0001) subgroups, relative to the MDX low pCR group. In a multivariable model, adjusted by clinical parameters (tumor size, node status, grade, age, and HER2 status), both the MDX intermediate pCR (OR: 2.82; 95% CI: 1.59-5.19, *p* = 0.0003) and MDX high pCR (OR: 11.42; 95% CI: 6.21-21.94, *p* < 0.0001) subgroups remained independent predictors of higher pCR rates (**Fig. 2b**). A qualitative comparison of pCR rates between the different treatment groups indicates that in the high pCR probability subgroup, the addition of carboplatin may have provided some incremental benefit, (68.6% vs 84.4%), but no additional benefit from the further addition of pembrolizumab was shown (84.4% vs 79.7%). In contrast, in the intermediate pCR probability subgroup, the incremental benefits of adding carboplatin and pembrolizumab were each quite apparent (increasing the probability of pCR from 31.5% to 45.0% and 61.9%, respectively). In the low pCR probability group, there was little difference in pCR rates between CTx+Cb+ICI+ and CTx+Cb+ICI− groups (30.4% vs 26.7%), but the pCR rate was nearly doubled in both groups compared to the CTx+Cb−ICI− group (16.0%).

**Figure 2.**
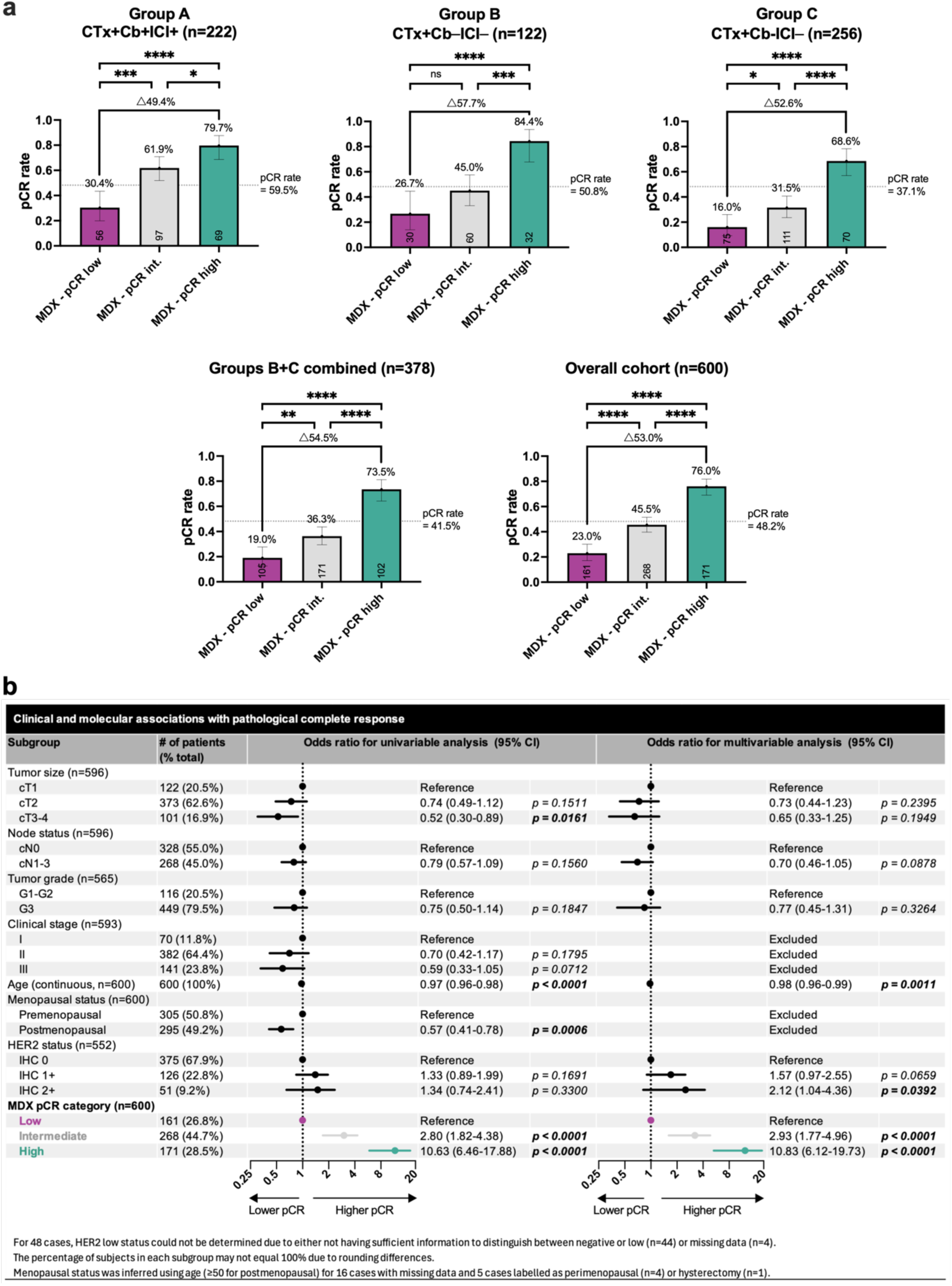
MDX pCR probability groups are associated with differential responses to neoadjuvant treatment. **a**) Bar charts show the pCR rates for patients classified in the MDX pCR low, intermediate, high probability groups in the CTx+Cb+ICI+ group, CTx+Cb+ICI−group, CTx+Cb−ICI− group, combined CTx+Cb+ICI− and CTx+Cb−ICI− group, and the overall cohort. The dotted lines denote the overall pCR rate of the arm irrespective of biomarker subgroup. Text above the bars show the pCR rates for each group. The connecting lines show the difference (delta Δ) in pCR rates between MDX pCR low or high groups. The numbers within each bar near the x-axis show the number of patients in each group. The error bars show the 95% confidence intervals (CI). **b**) Univariable and multivariable logistic regression illustrate the relationship between key clinical parameters and MDX pCR probability groups with pCR. Forest plots show the odds ratio point estimates and 95% CIs. Statistically significant (*p* < 0.05) comparisons are in bold text.

### Performance of Multiplex8+ pCR probability groups in clinical subgroups

We further investigated how MDX pCR probability groups stratify patients in different clinical subgroups (**Fig. 3a**). Patients classified in the MDX pCR high subgroup showed higher pCR rates in all clinical subgroups, with extraordinary performance in lower risk subgroups achieving an 86% (30/35) pCR rate in cT1 tumors (OR: 20.25; 95% CI: 6.36-76.83, *p* < 0.0001) and 91% (20/22) pCR rate in patients with stage I disease (OR: 37.50; 95% CI: 7.25-312.1, *p* < 0.0001). In general, the MDX pCR high subgroup also displayed statistically significant increased pCR rates in higher risk categories, but there was a trend towards lower pCR rates in larger tumors (76% in cT2 vs 65% in cT3-4 tumors). Patients with MDX pCR intermediate results also had higher odds ratios of achieving pCR, compared to those with MDX pCR low results in most clinical subgroups, including those with a higher tumor burden at initial diagnosis (e.g., cT3-4).

**Figure 3.**
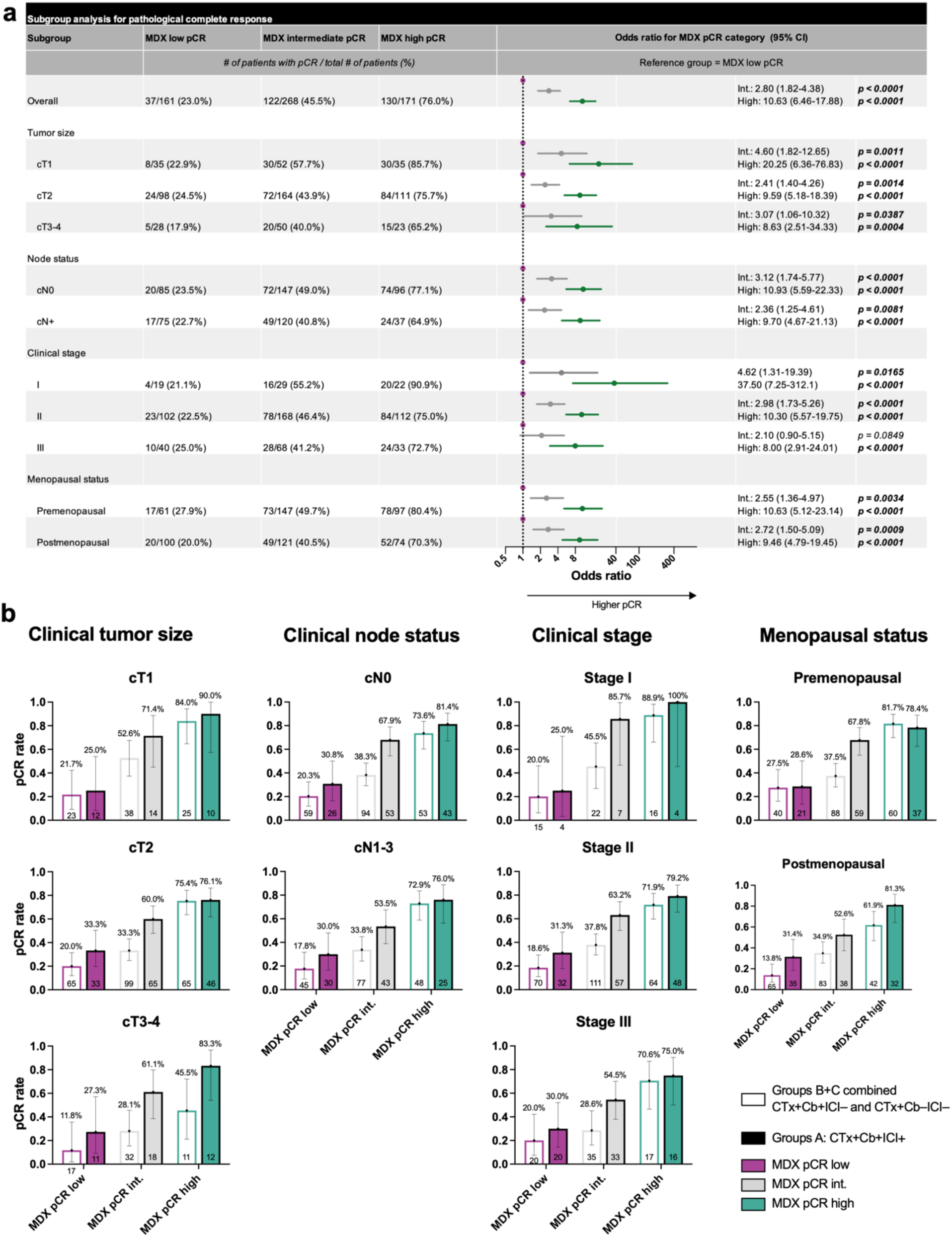
The MDX pCR probability groups have significantly different pCR rates in clinical subgroups. **a**) The table shows pCR rates between MDX pCR probability groups in clinical subgroups. Forest plots show the odds ratio point estimates and 95% confidence intervals (CI) of MDX pCR probability groups stratified by clinical subgroups obtained from univariable logistic regression. Statistically significant (*p* < 0.05) comparisons are in bold text. **b**) Bar charts show the pCR rates in the combined CTx+Cb+ICI− and CTx+Cb−ICI− group (open bars) and CTx+Cb+ICI+ group (solid bars) for patients classified in the MDX pCR low, intermediate, high probability groups. The text above the bars shows the pCR rates. The numbers within each bar or below the x-axis show the number of patients in each subgroup. The error bars show the 95% confidence intervals (CI).

The differences in pCR rates between MDX pCR probability groups in clinical subgroups remained consistent after splitting the overall cohort into group A and a combined chemotherapy group (i.e., Groups B and C; **Fig. 3b**). Notably, in some clinical subgroups (e.g. cT2, cN1-3, and premenopausal) the MDX pCR high group had equivalent or even higher pCR rates in the combined chemotherapy groups (B+C) compared to group A.

### The MDX-Risk signature is associated with RFS and OS

The MDX-Risk signature stratifies patients into MDX-low and MDX-high risk groups, with divergent RFS and OS. Irrespective of the treatment group, patients with an MDX-low risk result had better outcomes than those with MDX-high risk results, with 3-yr RFS and OS ranging from 89-94% and 98-100%, respectively, in all groups as well as the combined chemotherapy groups (B+C) and overall cohort (**Fig. 4a** and **Extended Data Fig. 4a**). In contrast, patients with MDX-high risk results had poorer outcomes with 3-yr RFS and OS ranging from 66-77% and 73-87%, respectively.

**Figure 4.**
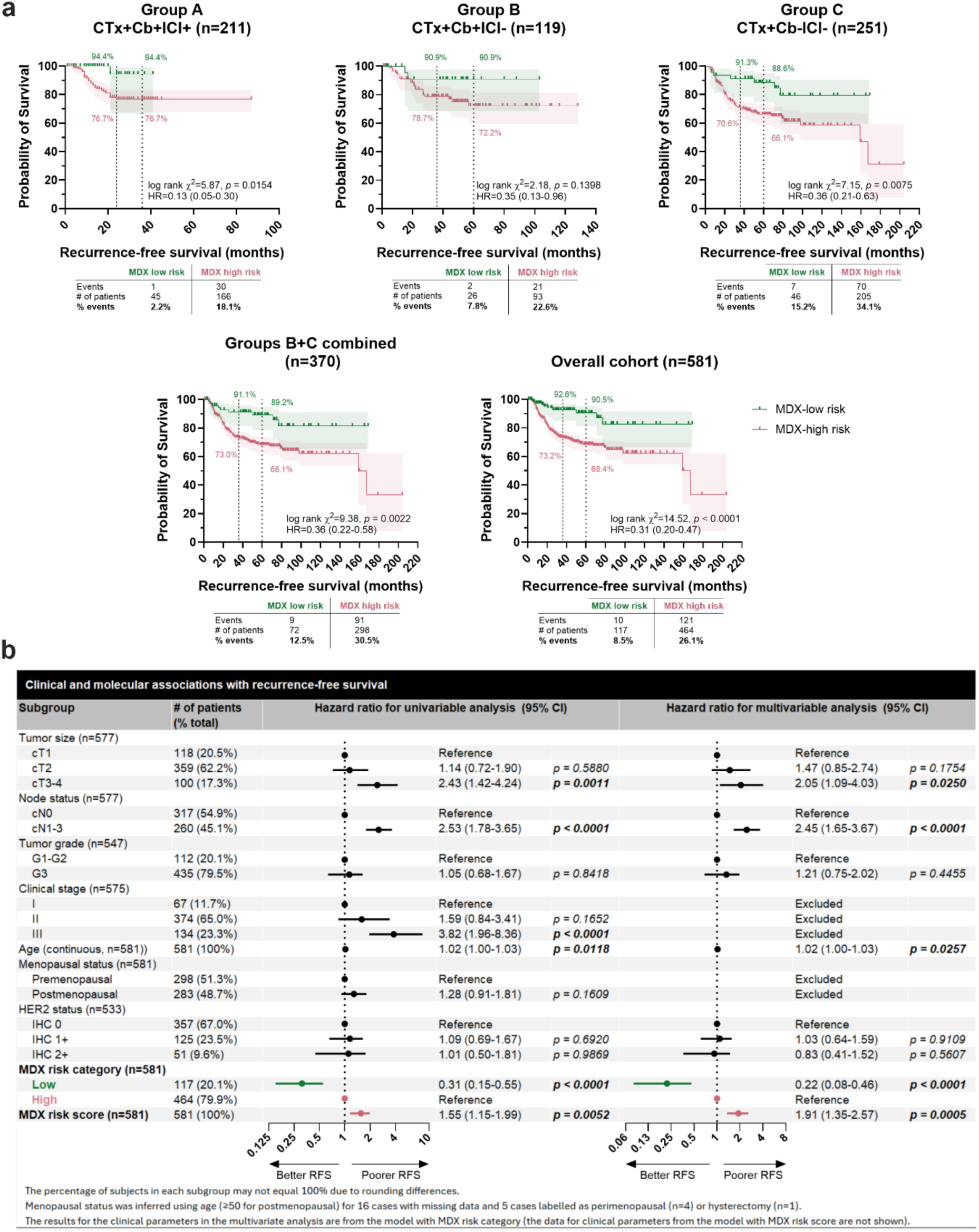
The MDX prognostic signature is associated with recurrence-free survival (RFS) in early TNBC. **a**) Kaplan-Meier curves show the probability of RFS in patients classified as either MDX high or low risk assessed in the CTx+Cb+ICI+ group, CTx+Cb+ICI− group, CTx+Cb−ICI−, combined CTx+Cb+ICI− and CTx+Cb−ICI− group, and the overall cohort. The dotted lines denote 2-yr and 3-yr RFS for the CTx+Cb+ICI+ group and 3-yr and 5-yr RFS for all other groups. The colored annotations show the probability of RFS for each risk group at the respective time points. The shaded areas show the 95% confidence interval (CI) of each curve. A logrank (Mantel-Cox) test was used to assess statistical differences between curves and the logrank method was used to derive hazard ratios. The table below each Kaplan-Meier curve shows the number of recurrence events, total patients (censored+ recurrence events), and the percentage of recurrence events. **b**) Univariable and multivariable Cox proportional hazards models illustrate the relationship between key clinical parameters and the MDX prognostic signature score (a continuous variable) and the corresponding MDX high and low risk groups with RFS. Forest plots show the hazard ratio point estimates and 95% CIs. Statistically significant (*p* < 0.05) comparisons are in bold text.

The 5-yr RFS in the overall cohort for MDX-high and MDX-low risk groups was 90.5% and 68.4%, respectively, which translated into large differences in 5-yr OS (96.5% and 76.8%). Univariable and multivariable Cox proportional hazard models revealed strong associations between the MDX-Risk signature, as a category or continuous score, and RFS (**Fig. 4b**) and OS (**Extended Data Fig. 4b**). MDX-low risk, relative to MDX-high risk, remained an independent prognostic marker in a multivariable model adjusted by known prognostic clinical parameters such as tumor size, node status, tumor grade, age, and HER2 status (low vs zero), with a robust 78% reduction in risk (HR: 0.22; 95% CI 0.08-0.46, *p* < 0.0001). Similarly, the continuous MDX-Risk signature score showed strong prognostic significance in a separate multivariable model adjusted by the same clinical parameters (HR: 1.91; 95% CI 1.35-2.57, *p* = 0.0005).

### Performance of the MDX-Risk signature in clinical subgroups

The MDX-Risk signature – expressed as either a category or continuous score – was strongly associated with RFS (**Fig. 5a**) and OS (**Extended Data Fig. 5a**) in most clinical subgroups. Patients with stage II disease showed the best stratification with only five recurrence events (5/81=6.2%; HR for RFS: 0.27; 95% CI: 0.09-0.59, *p* = 0.0005) and one death (1/82=1.2%; HR for OS: 0.09; 95% CI: 0.00-0.40, *p* =0.0002) in the MDX-low risk group compared to 68 recurrence events (68/293=23.2%) and 43 deaths (43/298=14.4%) in the MDX-high risk group. In contrast to the MDX pCR probability groups, the MDX-Risk groups and continuous score showed reduced performance in lower clinical risk groups (cT1 and stage I tumors); however, these subgroups had smaller sample sizes, fewer events, and consequently wider confidence intervals, thus warranting further investigation. These results in the overall cohort were consistent across clinical subgroups after stratifying into the combined chemotherapy groups (B and C) and group A (**Fig. 5b and Extended Data Fig. 5b**).

**Figure 5.**
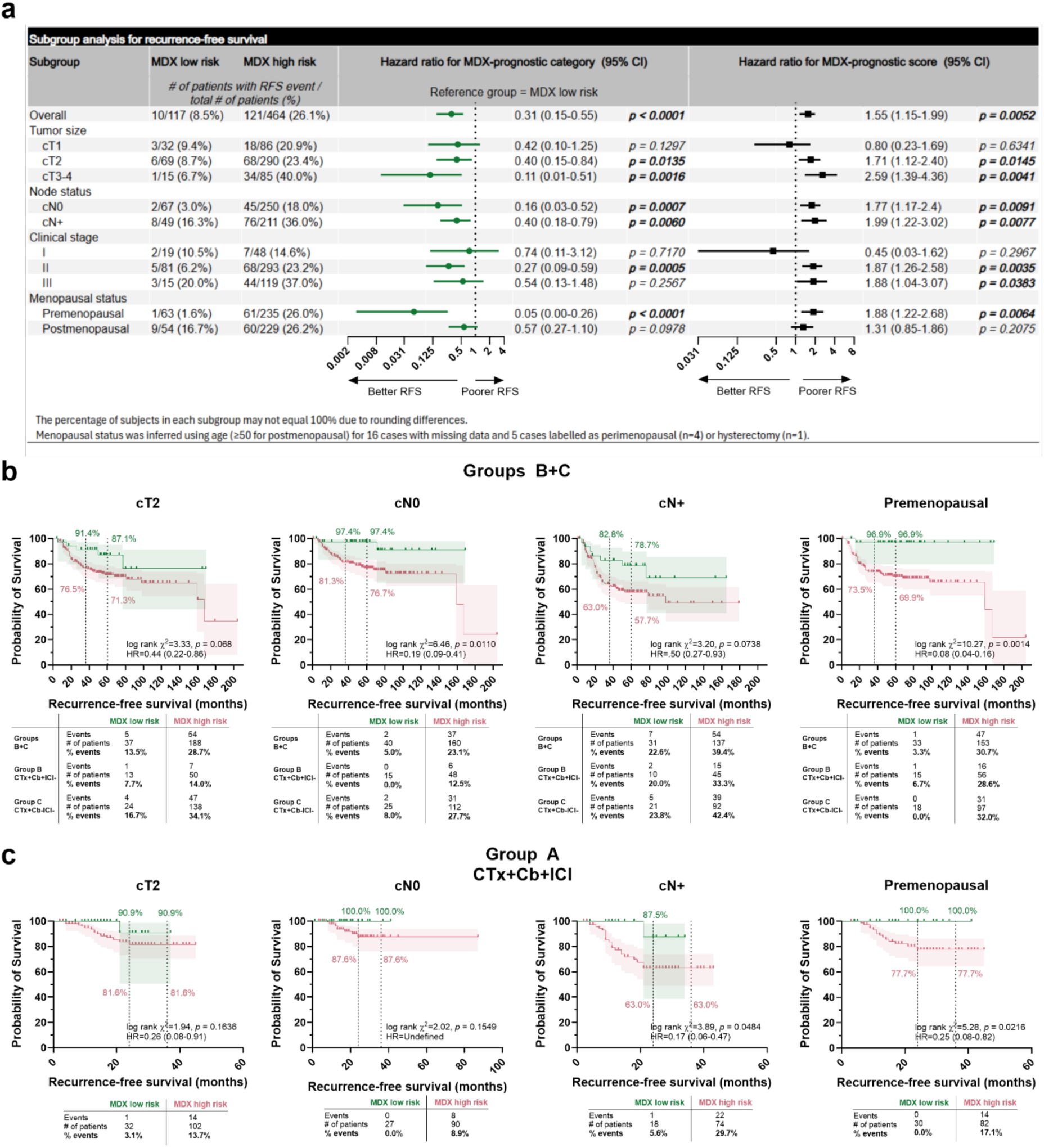
The MDX prognostic signature is strongly associated with recurrence-free survival (RFS) in key clinical subgroups. **a**) Univariable Cox proportional hazards models illustrate the relationship between the MDX high and low risk groups and corresponding MDX prognostic signature score (a continuous variable) with RFS in a subgroup analysis. Forest plots show the hazard ratio point estimates and 95% confidence interval (CI). Statistically significant (*p* < 0.05) comparisons are in bold text. **b-c**) Kaplan-Meier curves show the probability of RFS in patients classified as either MDX high or low risk assessed in the combined CTx+Cb+ICI− and CTx+Cb−ICI− group (**b**) and CTx+Cb+ICI+ group (**c**) for exemplary clinical subgroups. The dotted lines denote 3-yr and 5-yr RFS for the combined CTx+Cb+ICI− and CTx+Cb−ICI− group and 2-yr and 3-yr RFS for the CTx+Cb+ICI+ group. The colored annotations show the probability of RFS for each risk group at the respective time points. The shaded areas show the 95% confidence interval (CI) of each curve. A logrank (Mantel-Cox) was used to assess statistical differences between curves and the logrank method was used to derive hazard ratios. The table below each Kaplan-Meier curve shows the number of recurrence events, total patients (censored+recurrence events), and the percentage of recurrence events.

### Benchmarking MDX pCR and MDX-Risk signatures

We compared the performance of our MDX pCR probability groups and MDX-Risk signature with other biomarkers that have demonstrated predictive/prognostic potential in eTNBC, including: stromal tumor infiltrating lymphocytes (TILs) by pathologist assessment of H&E images; *CD274* (PD-L1) and *PDCD1* (PD-1) mRNA expression, the T-cell inflamed gene expression signature (T-cell GEP)^40,41^, the response predictive subtypes 5 (RPS-5) classification scheme developed in I-SPY-2^34^, and a combined 4-gene tumor cell proliferation signature and 10-gene core immune group (CIG) module^42^. These various gene expression profiling assessments were bioinformatically applied to the same LCM tumor-enriched RNAseq data used for our MDX analyses. All biomarkers, expressed as either categories or continuous scores, were associated with PCR, RFS, and OS in both univariable and multivariable Cox models, with the multivariable model containing the biomarker and TILs (**Extended Data Fig. 6a-c** and **Extended Data Fig. 7a-c** for continuous scores). While most showed similar performance, the MDX high pCR probability group and the MDX low risk group had the highest odds ratio and lowest hazard ratio, respectively in both univariable and multivariable models. Notably, in the multivariable models with TILs, the MDX-Risk signature was the only example where both the biomarker signature and TILs remained independently associated with RFS/OS. This suggests that while other signatures capture most of the information provided by TILS, the MDX-Risk signature provide complementary information that may synergize with TILs. A heatmap showing the relationship between the biomarkers and clinical outcomes is shown in **Extended Data Fig. 7d**.

### Benchmarking the performance of biomarker signatures in specimens that did or did not undergo laser capture microdissection

Since inclusion of non-tumoral tissue (e.g., healthy, DCIS, stromal admixture) may confound gene expression signatures, we compared 509 samples that were sequenced with or without LCM. While LCM modestly improved the individual MDX-ICI (2.9% gain in accuracy) and MDX-CTx (1.2% gain in accuracy) signatures (**Extended Data Fig. 8a-b**), this translated into a greater pCR rate difference between pCR high vs low probability groups in LCM samples (53.1% difference: OR: 10.66; 95% CI: 5.94-18.95, *p* < 0.0001) compared to no LCM (43.8% difference: OR: 6.58; 95% CI: 3.94-11.16, *p* < 0.0001; **Extended Data Fig. 8c**). Prior LCM also improved stratification of MDX low and high-risk groups (RFS HR for LCM: 0.34; 95% CI: 0.22-0.54 vs RFS HR for No LCM: 0.48; 95% CI: 0.30-0.75; **Extended Data Fig. 8d**). LCM, compared to No LCM, had a profound impact on TNBC subtyping, with 43.7% of samples changing subtypes, driven by an increased proportion of samples classified in the immunomodulatory (IM) and mesenchymal stem-cell like (MSL) subtypes in bulk processed samples (**Extended Data Fig. 8e**).

## Discussion

The treatment of early triple-negative breast cancer has shifted decisively toward intensification, with carboplatin and pembrolizumab now added to conventional chemotherapy for high-risk disease. This escalation has improved pathologic response and survival, but it applies the most toxic regimen uniformly to a biologically heterogeneous population — exposing patients who would respond to less intensive therapy to avoidable toxicity, while offering no means to identify those for whom even intensified treatment may be insufficient. Resolving this tension requires biomarkers that are both predictive, distinguishing who benefits from a given therapy, and prognostic, identifying who remains at risk despite treatment. Such markers would enable rational de-escalation for patients likely to respond to less intensive regimens and escalation for those at high residual risk, aligning treatment intensity with individual biology rather than population-level averages.

To address this need, we developed three gene signatures on a large, internationally diverse corpus of 1,063 patients diagnosed with TNBC spanning 28 studies and validated them in a real-world, multicenter, retrospective cohort of 602 samples from 590 patients with eTNBC. In the present study, we found the MDX-CTx and MDX-ICI response signatures, which were trained on clinical cohorts treated with either chemotherapy or chemoimmunotherapy, respectively, were prognostic for improved pCR rates in all treatment groups and not specific to treatment type, highlighting that common biological mechanisms may underlie the response to both chemotherapy and immunotherapy. This is consistent with biomarker analyses of KN-522^43^, GeparNuevo^44^, and NeoTRIP^45^, which reported substantial overlap in the baseline biological features associated with neoadjuvant response, especially those encompassing immune-activated tumors (e.g., PD-L1, TILs, immune gene signatures). We further demonstrated these two signatures synergized to form three groups – low, intermediate, and high – with increasing likelihood of achieving a pCR. The MDX high pCR group had remarkable responses irrespective of treatment, demonstrating minimal added benefit of ICIs (79.7% pCR rate in CTx+Cb+ICI+ vs 84.4% in CTx+Cb+ICI−. In contrast, the MDX low and intermediate pCR groups showed more drastic improvements in pCR rates with the addition of Cb or both Cb and pembrolizumab.

The MDX-Risk signature, trained to identify risk of early recurrence, stratified patients into high and low risk groups with significantly different longitudinal outcomes, with patients classified as low-risk having 3-yr RFS and OS of 92.6% and 98.0%, respectively, with only marginal differences between patients treated with CTx or ICI. All signatures provided independent predictive/prognostic information in multivariate models with clinical variables like tumor size and node status and performed consistently across key clinical subgroups. In the **Supplementary information**, we also illustrate a hypothesis-generating framework where the MDX-Risk signature further stratifies the MDX intermediate pCR group, creating an optimized binary clinical outcome stratification framework associated with either a Favorable Response & Prognosis Profile (FRPP, MDX high pCR + MDX intermediate pCR/MDX-low risk patients) or Adverse Response & Prognosis Profile (ARPP, MDX low pCR + MDX intermediate pCR/MDX-high risk). The Response & Prognosis Profiles (RPP) framework integrates with clinical parameters, existing biomarkers like TILs, and neoadjuvant response to illuminate subsets of patients who could be considered for treatment optimization paradigms in future prospective clinical trials.

The pivotal KN-522 trial ensconced pembrolizumab combined with taxanes and carboplatin followed by anthracyclines and cyclophosphamide as the new SoC for high-risk eTNBC, demonstrating gains in pCR rates^7^ and prolonged event-free survival (EFS)^8^ and OS^9^. However, the 4.9% overall survival gain means that for every 100 patients treated with pembrolizumab only five derive additional benefit over standard chemotherapy. The challenge of balancing these gains with the KN-522 regimen’s significant potential for toxicity, creates urgency to develop novel biomarkers and tools to identify those who are most likely to benefit from treatment intensification, while sparing others from avoidable toxicity. The most common de-escalation approach is to remove anthracyclines, due to their potential cardiotoxicity, in favor of a taxane/carboplatin backbone that can be combined with immunotherapies like pembrolizumab. Anthracycline-free chemotherapy and chemoimmunotherapy regimens have shown promising results in phase II trials like WSG-ADAPT-TN^46^, NeoCART^47^, NeoPACT^48^, and NeoSTOP^49^, reporting competitive and even superior pCR rates and long-term survival compared to anthracycline-based regimens. In a pooled analysis of two prospective eTNBC cohorts receiving an anthracycline-free regimen of docetaxel + carboplatin, BRCA status, which is linked to platinum sensitivity was not associated with pCR^50^, thus highlighting the need for biomarkers.

The MDX pCR probability groups and MDX-Risk signature could be used to select patients for de-escalation in future prospective trials. Patients in the high MDX pCR probability group had exceptional responses with pCR rates of nearly 85% in the CTx+Cb+ICI− group thus revealing a group of patients that may have excellent outcomes on less intense treatment regimens. Likewise, patients with a low MDX-Risk signature had 5-year RFS of 90.5% and OS of 96.5% that surpassed the outcomes of the pembrolizumab-chemotherapy group in the KN-522 trial (5-year RFS of 81.2% and OS of 86.6%)^9^. Since these gains were evident in the chemotherapy only groups, these patients may be less likely to derive an absolute benefit from additional pembrolizumab. Our RPP framework also showed how these signatures could synergize with clinical burden to further refine patient selection.

Patients classified in the high MDX pCR probability group, MDX-low risk, or in the FRPP group, especially those with lower clinical burden, may constitute an ideal candidate population for future de-escalation trials exploring anthracycline-free, carboplatin-based chemo- or chemo-immunotherapy regimens or lower intensity pembrolizumab regimens.

Despite a growing treatment armamentarium in eTNBC, many patients have intrinsic resistance to neoadjuvant chemo or chemoimmunotherapy. In KN-522, 35% of patients treated with pembrolizumab and 50% of patients treated in the control arm had residual disease (RD), and consequently a poor prognosis. A variety of emerging therapies such as ADCs, bi-specific antibodies, and combination therapies are being investigated in eTNBC in both neoadjuvant and adjuvant settings^3,51^. Our diagnostic assay could help highlight these patients by assessing baseline neoadjuvant treatment sensitivity and prognosis. We found patients classified as MDX pCR low have extremely low pCR rates (ranging from 16.0% in CTx+Cb+ICI− to 30.4% in CTx+Cb+ICI+), approximately half of the average pCR rate in each treatment group. Patients with MDX-high risk have poor outcomes irrespective of treatment (ranging from 3-yr RFS of 76.7%, 78.7%, and 70.6% for groups A, B, and C, respectively). Similarly, patients with an ARPP and either higher clinical burden (3-yr RFS of 54.4%), low TILs (3-yr RFS of 53.2%), or RD (3-yr RFS of 44.6%) had the poorest outcomes. In a curative-intent setting, identifying these patients upfront is critical as they could be enrolled in clinical trials investigating novel therapies, rather than wasting an opportunity by administering treatment that has a low probability of efficacy.

The MDX pCR probability groups, MDX-Risk signature, and RPP framework could also be combined with early mid-treatment monitoring, either through imaging (e.g., magnetic resonance imaging, MRI) or ctDNA. For example, upon evidence of a radiologic complete response or cleared ctDNA, patients with MDX-high pCR probability, MDX low risk, or FRPP could potentially bypass more toxic blocks of chemotherapy and proceed directly to surgery. A similar approach is being tested in the adaptively randomized I-SPY 2.2. trial (NCT01042379). Alternatively, if imaging indicates disease progression or ctDNA rises, patients with MDX-low pCR probability, MDX high risk, or ARPP could rapidly be detected as non-responders and guided to alternative therapies in clinical trials.

We observed that certain MDX signatures (i.e., MDX-CTx, MDX-Risk, and RPP framework) may synergize with TILs, providing an enticing way to integrate Multiplex8+ with other novel biomarkers and existing clinical workflows. Multivariate models revealed enhanced stratification when the MDX-CTx and MDX-Risk signatures were combined with TILs, providing clear rationale for their combination. In our hypothesis-generating RPP framework, patients with a FRPP with high TILs had better outcomes than those with low TILs. In contrast, patients with an ARPP had poor outcomes irrespective of TILs, suggesting that TILs alone may be insufficient to identify patients for de-escalation. This is germane to ongoing and future trials where TILs are used as integral biomarkers for patient selection to de-escalate treatment for patients with stage I-III disease in the neoadjuvant (NeoTRACT^52^, DespaTIL^53^, cohort C of BELLINI^54^) or adjuvant (OPTImal^55^, ETNA^56^) setting. Our findings warrant further investigation in future prospective trials and translational analyses of existing trials by integrated transcriptomic profiling with TILs to provide more granular stratification of patients for de-escalation.

Neoadjuvant response can help tailor adjuvant therapy, with patients who achieve pCR considered for de-escalation and those with residual disease for escalation. Because KN-522 did not randomize postoperative pembrolizumab, the relative contributions of its neoadjuvant and adjuvant components remain uncertain, prompting questions about the necessity of adjuvant therapy. Notably, GeparNuevo^57^ and I-SPY2^58^, which administered immunotherapy only in the neoadjuvant setting, reported 3-year EFS comparable to KN-522, despite omitting adjuvant immunotherapy. The ongoing phase III OptimICE-PCR trial (NCT05812807) aims to resolve this issue by randomizing patients with eTNBC and pCR to adjuvant pembrolizumab or observation. Notably, our findings that MDX FRPP/ARPP groups synergize with clinical stage, and neoadjuvant response may help identify patients suitable for adjuvant de-escalation. Patients with a MDX FRPP and pCR had exceptional outcomes with >97% RFS/OS; however, perhaps more importantly, patients with a MDX ARPP and pCR were more susceptible to having a recurrence event (11.1% vs 5.5%) or death (5.9% vs 2.2%). This suggests pCR alone is insufficient to safely de-escalate, but rather combining biomarker signatures with pCR may provide a more reliable approach to navigating this important treatment decision.

Our study has some limitations. Due to the retrospective study design, our cohort may be confounded by selection biases such as the different treatment groups being the SoC during different historical periods. This, together with the lack of randomization, precludes robust statistical comparisons of MDX signature performance between treatment groups and limits the interpretation of long-term follow up data for the CTx+Cb+ICI+ group. However, we obtained nearly 5-yrs of follow up in the CTx+Cb+ICI− and CTx+Cb−ICI− groups and the MDX signatures showed similar performance between the combined CTx+Cb+ICI− and CTx+Cb−ICI− group and CTx+Cb+ICI+ group, suggesting that long-term outcomes may extrapolate to the CTx+Cb+ICI+ group.

Altogether, we validated three gene signatures that track baseline neoadjuvant treatment sensitivity and long-term risk of recurrence in a real-world, multicenter, retrospective study on 590 patients (602 samples) with eTNBC treated with either chemo or chemoimmunotherapy. These signatures outperformed common biomarkers and clinical parameters, and, moreover, can be combined and synergize with both clinical parameters and neoadjuvant response to identify patients that could undergo treatment optimization through de-escalation or escalation strategies in both neoadjuvant and adjuvant treatment settings.

## Methods

### A real-world, multicenter, retrospective cohort of patients with eTNBC

We conducted a real-world, multicenter, retrospective study on patients treated with either neoadjuvant chemotherapy (with or without carboplatin) or neoadjuvant chemoimmunotherapy (e.g., the KN-522 regimen). The study included 590 patients (602 samples) from nine sites across eight countries undergoing routine standard of care neoadjuvant treatment and were divided into three treatment groups: the KEYNOTE-522 regimen (group A, CTx+Cb+ICI+, regimens that included: pembrolizumab plus paclitaxel plus carboplatin (Cb) followed or preceded by pembrolizumab plus doxorubicin (Adriamycin, A) or epirubicin (E) plus cyclophosphamide (C; n=219); pembrolizumab plus paclitaxel followed or preceded by pembrolizumab plus AC or EC (n=5)), CTx with Cb (group B, CTx+Cb+ICI−, regimens that included: AC or EC followed or preceded by taxanes (T, paclitaxel or docetaxel) plus carboplatin (Cb, n=101); T plus Cb (n=15); AC followed by cisplatin (Cis; n=5); and AC followed by T plus Cis (n=1)), and CTx without Cb (group C, CTx+Cb−ICI−, regimens that included: AC or EC followed or preceded by T (n=217); AC or EC (n=12); TC (n=11); 5-fluorouracil (F) plus EC followed by docetaxel (FEC-D, n=8); T (n=6); FEC (n=1); and E (n=1)). Male and Female patients were included in the study if they were ≥ 18 years old at the time of informed consent, had histologically confirmed hormone receptor (HR) negative (<1%) or HR-low positive (defined as ER and/or PR 1-10% of positive cells by IHC) and HER2− breast cancer (IHC 0, 1+, and 2+ with equivocal results confirmed with DNA-FISH/SISH/ISH as non-amplified considered as negative) according to ASCO/CAP guidelines, and had received (in the past or during enrollment) neoadjuvant treatment. Patients were excluded from the study if they had a known second primary malignancy or were missing both pCR and RFS data. Informed consent from each patient was obtained from the respective site, and no tissues were processed without informed consent.

The primary endpoint of the study was pCR, defined according to the NeoSTEEP^59^ standardized definition as the absence of residual invasive cancer in the breast and axillary lymph nodes [ypT0/Tis ypN0]) at the time of definitive surgery. The secondary endpoint was RFS, defined as the time from date of neoadjuvant treatment initiation to date of first event of local invasive breast recurrence, regional invasive recurrence, distant recurrence, or death of any cause. OS, an exploratory endpoint, was defined as the time from date of neoadjuvant treatment initiation until death of any cause. For statistical analyses and data visualization, samples from patients with multiple biopsies (n=12) were treated as independent samples.

Participating sites also provided a standardized clinicopathological data table with information linked to each sample including age, clinical and pathological tumor size, node status, histological grade and type, neoadjuvant and adjuvant therapy history, genomic testing results, neoadjuvant response (pCR), and RFS/OS. The clinical data followed the Biospecimen Reporting for Improved Study Quality (BRISQ) criteria^60^ and was used to perform association analyses with molecular data.

### Study plan

The Ethics Committee of the Bratislava Self-Governing Region gave ethical approval for this work (Ref. No. 05320/2020/HF). In addition, the study protocol was approved by local ethical committees for each of the participating sites that provided anonymized patient specimens and clinical data. Each site obtained informed consent from the patients for inclusion in the study.

All sites, except Biobank Graz and Sunnybrook, sent FFPE blocks to MultiplexDX where biopsies were sectioned on a microtome at 5 μm, placed on positive charged glass slides or functionalized PEN membrane slides, which were used for hematoxylin and eosin (H&E) staining, multiplexed RNA-FISH, and a rapid cresyl violet staining for laser capture microdissection. Due to restrictions on sending blocks externally, Biobank Graz and Sunnybrook sectioned the blocks according to a Standard Operating Protocol with onsite supervision (Graz) or prior training (Sunnybrook) by MultiplexDX staff to ensure reproducibility.

Multiplexed RNA-FISH (Advanced Cell Diagnostics RNAscope™ Multiplex Fluorescent V2 Assay) was used to detect the transcripts for progesterone (*PGR*), estrogen (*ESR1*), and HER2 (*ERBB2*) receptors, and the proliferation marker Ki67 (*MKI67*). Whole slide images (obtained from an Akoya Phenoimager HT 2.0) of RNA-FISH and H&E were used to determine any biomarker or histological heterogeneity and to guide appropriate regions of interest (ROI) for laser capture microdissection. ROIs were selected based on pathologist-guided annotations on H&E scans. If the sample was heterogenous in biomarker expression, the RNA-FISH scan was jointly used to guide selection of ROIs. Areas of up to 39.92 mm^2^ were dissected from up to 6 sections, with additional sections used only until a minimum combined area of 10 mm^2^ was reached. ROIs were collected from an adjacent section stained with a rapid cresyl violet solution to preserve RNA integrity and stored in a lysis buffer until further automated RNA extraction using a KingFisher instrument (Thermofisher MagMAX™ FFPE DNA/RNA Ultra Kit). RNA quantity and quality were assessed using a Qubit 4 Fluorometer and Agilent 4150 TapeStation and then total RNA sequencing libraries (SMART-Seq Total RNA Library Prep with ZapR Depletion (with UMIs)) were prepared from specimens that had sufficient quantity (≥ 40 pg) and quality (DV200 values > 5%). Successfully prepared libraries (≥ 0.5 ng/μl) were pooled, spiked with 10% PhiX, and sequenced on an Illumina NovaSeq 6000 (paired-end reads 2 x 100 bp; approximately 50-100 M reads per sample). Sequencing data was analyzed using a standard bioinformatic pipeline (see below).

Patient specimens were processed in batches according to a stratified randomization approach, ensuring that each batch comprised a representative sampling of treatment groups (i.e., KN-522 regimen and chemotherapy with and without carboplatin) and neoadjuvant response (pCR vs residual disease). Batches consisted of at least 6 samples (6-24 samples depending on the laboratory technique) and were nested into four layers of batches that follow a sequential order: 1) Microtome, RNA-FISH, and LCM (6-16 samples per batch, including samples with more than one sample dissected by LCM), 2) automated RNA isolation (7-12 samples), 3) RNA-SEQ library preparation (7-38 samples), and 4) RNA-sequencing on an Illumina NovaSeq6000 (up to 180 samples per run). Each batch was processed by an individual researcher and batches earlier in the workflow were maintained in later batches.

### Tissue processing and H&E staining

At least eleven 5 μm sections from FFPE breast cancer specimens were obtained using a Leica Histocore Multicut microtome. Consecutive sections were collected in the following order: 1 section on a glass slide for backup H&E, RNA-FISH, or other molecular analysis, 1 section on a glass slide for H&E, 2 sections on a functionalized PEN membrane slide for LCM (both sections placed on the same slide), 2 sections on a second functionalized PEN membrane slide for LCM (both sections placed on the same slide), 1 section on a glass slide for RNA-FISH, 1 section on a glass slide for RNA-FISH, and two sections taken as scrolls and placed directly into separate 1.5 mL microcentrifuge tubes and frozen at –20 °C for later RNA extraction and RNA sequencing. To confirm the presence of invasive breast cancer, one section was stained using H&E, cover slipped and subjected to whole-slide scanning. Notably, the H&E-stained section was adjacent to the PEN membrane slide used for LCM to ensure comparable tissue morphology between the annotated slide and the slide used for microdissection.

### Multiplexed RNA-FISH

The Advanced Cell Diagnostics RNAscope™ Multiplex Fluorescent V2 Assay was used to detect *PGR*, *ESR1*, *ERBB2*, and *MKI67* following the manufacturer’s instructions. Each marker was visualized using Akoya Opal 690, 620, 520, and 570 fluorophores, respectively. This approach allowed us to map tumor tissue heterogeneity and identify ROIs for LCM which was then used to isolate key tumor-specific ROIs, while excluding surrounding healthy tissue, stroma, and adipose cells that could confound gene expression analysis.

### Whole-slide imaging, image annotation, and image analysis

Following H&E and RNA-FISH staining, whole-slide brightfield and fluorescence images were acquired using Akoya Vectra Polaris (Phenoimager HT 2.0) imaging system with a 20x objective lens (0.5 µm/pixel resolution) and standard Akoya MOTiF™ multispectral imaging filters. The H&E whole slide scans were annotated by a trained researcher and subsequently reviewed by a board-certified pathologist (D.A.E), using the open-source program QuPath v0.5.1 (https://qupath.github.io/) to identify invasive breast cancer, with color-coded annotations distinguishing histological subtypes when multiple were present in a specimen. The RNA-FISH whole slide scans were annotated by a trained researcher using Akoya Phenochart™ software v1.1.0 to qualitatively assess the intensity and distribution of fluorescent signals corresponding to *ESR1, PGR, ERBB2*, and *MKI67* markers. Based on the H&E and RNA-FISH annotations, regions of invasive carcinoma were selected for LCM by a trained researcher. In specimens containing multiple histological subtypes, each region of interest was separately subjected to LCM and downstream analyses (Supplementary Methods).

For RNA-FISH image analysis, microdissected ROIs were stamped on the whole slide scans and analyzed for biomarker signals using Akoya InForm® software v2.6.0. Depending on the size of the dissected area, 1-3 stamped regions were analyzed. The analysis included: (1) spectral unmixing and autofluorescence isolation using a synthetic spectral library; (2) machine learning-based tissue segmentation (tumor vs. stroma) and cell segmentation into nuclear and cytoplasmic compartments; and (3) biomarker expression scoring. Average fluorescence intensity for each marker was quantified within the tumor segment of the image. The researcher performing the analysis was blinded to IHC results and clinicopathological data.

Stromal tumor-infiltrating lymphocytes (TILs) were assessed in H&E sections by a board-certified anatomic pathologist (D.A.E.) following recommendations of the International Immuno-Oncology Biomarker Working Group on Breast Cancer^61^.

### Laser capture microdissection

Established protocols from Leica were followed for conducting LCM in a manner that maintained RNA integrity. This included rapid cresyl violet staining, limiting dissection times to under 1 hour per sample, and taking precautionary measures to preserve RNA integrity. Regions for microdissection were identified by comparing the annotated H&E and RNA-FISH images with the adjacent cresyl violet-stained section. Approximately 10 mm² of tissue was dissected per sample to ensure sufficient material for RNA extraction. For specimens with limited tumor area, we conducted LCM on multiple PEN membrane slides to obtain the required material. After LCM, specimens were snap-frozen using 96% denatured ethanol and dry ice and stored in a -80°C freezer.

### RNA isolation and quality control

Total RNA was isolated using the MagMAX™ Ultra Kit (Thermo Fisher Scientific) on a KingFisher™ Duo Prime automated purification system (Thermo Fisher Scientific) according to the manufacturer’s instructions. Briefly, samples were lysed in protease solution for 60 minutes at 55 °C, followed by incubation for 15 minutes at 90 °C. The samples were then transferred to a pre-filled KingFisher deep-well 96-well plate (with wash buffers and DNase solution according to the given instructions) and mixed with magnetic beads. The KingFisher Duo Prime instrument automated magnetic bead binding, DNase treatment, washing, and elution steps using the manufacturer’s standard protocol. Purified RNA was then transferred from the elution strip to 1.5 mL tubes. RNA quantity was measured using the Qubit™ RNA HS assay Kit with a Qubit 4 Fluorometer and RNA integrity (DV200) using the Agilent High Sensitivity RNA ScreenTape System with an Agilent 4150 TapeStation. Following quality control, samples were snap-frozen using 96% denatured ethanol and dry ice and stored at −80 °C.

### RNA library preparation and sequencing

Total RNA-SEQ libraries were prepared using the SMART-Seq Total RNA Library Prep with ZapR Depletion (with UMIs) kit following the manufacturer’s instructions. After library preparation, the quantity and fragment size range of each library were assessed using both the Qubit 1x dsDNA HS kit with a Qubit 4 Fluorometer, and the Agilent High Sensitivity D1000 ScreenTape assay on an Agilent 4150 TapeStation.

Individual libraries containing sufficient concentration (≥0.5 ng/μl) were pooled. The average size and molarity were accessed using the Agilent High Sensitivity D1000 ScreenTape assay on an Agilent 4150 TapeStation. Fragments ranged between 200 and1000 bp, with an average of∼350 bp. Library pools were sequenced on an Illumina NovaSeq 6000 using SP, S1, S2, or S4 flow cells depending on pool size. Pooled libraries were spiked with 10% PhiX as recommended by both Illumina and Takara for low-complexity libraries. Paired-end sequencing (2 × 100 bp) was conducted with the aim of obtaining approximately 100 million reads per sample.

Raw sequencing data was checked for quality using MultiQC tool, followed by adapter trimming (Cutadapt), alignment to the latest human genome build (STAR), deduplication of unique molecular indices (UMI-tools), and then quantification of gene/transcript abundances across RNA biotypes. This will result in a gene matrix table that used for downstream analyses.

### Cohort descriptive statistics

Standard descriptive statistics were used to summarize sample characteristics for all eligible patients that meet the inclusion/exclusion criteria and yielded specimens that met the quality control criteria for processing. Descriptives were stratified by treatment group and presented as either median or mean, with the percentages of samples or ranges represented in parentheses.

### Statistical analyses of main endpoints

Fisher’s Exact tests (2x2, 2x3, and 2x4 contingency tables) were used to compare pCR rates among biomarker subgroups (e.g., MDX-CTx + vs −, MDX-ICI + vs −, MDX-Risk high vs low, MDX pCR probability groups, etc.), compute odds ratios, and test for significance followed by post hoc comparisons using Holm-Bonferroni to correct for multiple testing when necessary. The 95% confidence intervals (CIs) shown in the bar charts illustrating the proportion of patients with a pCR were derived using the modified Wald method^62^. Logistic regression was used to associate MDX signatures, expressed as either categorical or continuous variables, with the binary outcome (pCR or RD) in univariable and multivariable analyses, clinical subgroups, and benchmarking.

Kaplan-Meier (KM) curves were used to illustrate data involving either RFS or OS, with logrank (Mantel-Cox) tests used to determine whether two or more curves were significantly different and to derive hazard ratios and 95% CIs. Cox proportional hazards models were used to associate MDX signatures, expressed as either categorical or continuous variables, with either RFS or OS in univariable and multivariable analyses, clinical subgroups, and benchmarking.

One-way ANOVA followed by Tukey’s post hoc multiple comparisons test was used to compare MDX-Risk signature scores within pCR probability groups. Forest plots were used to illustrate the odds ratio or hazard ratio point estimates, 95% CIs, and p values for the logistic regression or Cox proportional hazards models, respectively. For all statistical analyses, the significance level was set at alpha of 0.05 (for both adjusted and unadjusted p-values).

## Data Availability

RNA-sequencing data, de-identified clinicopathological data table, derived prognostic/gene signatures, digitized whole slide images of H&E (with and without pathology annotations), digitized whole slide images and analyzed region of interest from multiplexed RNA-FISH cannot be publicly shared due to patient privacy and existing material and data transfer agreements between MultiplexDX and participating biobanks and commercial companies. Qualified researchers may apply for access to these data through the MultiplexDX Data Access Committee (DAC) by sending an initial request to the lead corresponding author (P.Č.) or the following email address. Then the qualified researcher would submit a brief research proposal and a standard form describing the project, data/materials requested, applicable ethics, and purpose. Requests will be reviewed and discussed by the DAC based on scientific merit, existing collaborations, and commercial agreements. The time frame of response to an initial request is about 1-2 months. After approval, the parties will agree on the conditions of a data access/sharing agreement and restrictions of use, which may increase the total time frame to around 6 months. Alternatively, qualified researchers may directly contact the nine sites that retrieved and supplied the diagnostic core biopsies and linked clinicopathological data.

Regarding external datasets, to develop a chemotherapy response signature, we used GSE205568 (https://doi.org/10.17605/OSF.IO/ZT8CU), which is compendium of 23 microarray gene-expression studies comprising 706 TNBC samples treated with neoadjuvant chemotherapy. For the development of the immunotherapy response signature, we used the pembrolizumab (n=29, GSE194040, 10.1016/j.ccell.2022.05.005) and the durvalumab plus olaparib (n=21, GSE173839, 10.1016/j.ccell.2021.05.009) arms of I-SPY2 study. For the MDX-Risk signature, we used the TNBC subset of MDX-BRCA cohort as a training dataset (n=159, GSE283522, 10.1038/s41467-024-55583-2) and TCGA-BRCA as a validation dataset (n=148, available at: https://portal.gdc.cancer.gov/, 10.1016/j.cell.2018.03.022, https://doi.org/10.1038/nature11412).

## Code Availability

All software used for data collection, analysis, and bioinformatics was either open source or commercially available as documented in the **Methods**, **Supplementary Information**, and **Reporting Summary**; therefore, no new source code was generated in this paper.

## Acknowledgements

We gratefully acknowledge Biobank Graz of the Medical University of Graz, Austria and PATH Biobank (Munich, Germany), Brno Masaryk Memorial Cancer Institute, St. Anne’s University Hospital in Brno, The European Interbalkan Medical Center (Thessaloniki), François Baclesse Center (Caen), Universitätsklinikum Leipzig, Medicyt Košice, UNILABS Košice, UNLP Košice, Sunnybrook Health Science Centre (Toronto), Hospital del Mar Research Institute (Barcelona) for providing the FFPE breast cancer specimens and associated clinicopathological data; We want to particularly acknowledge the patients and the Biobank MARBiobanc integrated in the ISCIII National Biomodels and Biobanks Platform for their collaboration; Dorota Adamska and other staff at the Centre for New Technologies, University of Warsaw, Genomics Core Facility for providing exceptional core sequencing services; MUDr. Tomáš Sieber and MUDr. Andrea Cipková, MPH. from the East Slovak Oncology Institute in Košice (Východoslovenský onkologický ústav a.s.) for consultations and feedback on patient follow-up data.

This project was partially funded by the EU NextGenerationEU through the Recovery and Resilience Plan for Slovakia under the project No. 09I01-03-V01-00003 (granted by the Ministry of Economy of the Slovak Republic) awarded to MultiplexDX s.r.o. (Pavol Čekan as PI), project No. 09I03-03-V03-00101 (granted by the Research and Innovation Authority (VAIA) of the Slovak Republic) awarded to MultiplexDX s.r.o. (Evan D. Paul as PI), as well as project PK 1/2025 under the project No. 250/2025 granted by Deputy Prime Minister of the Slovak Republic for the Recovery Plan and the Knowledge Economy awarded to MultiplexDX s.r.o. (Pavol Čekan as PI). RNA-sequencing data was obtained using the computational resources funded by Project 101083419 — SKAI-eDIH — DIGITAL-2021-EDIH-01. L.C., J. A. and S.Se. acknowledge support from CIBERONC CB16/12/00241, PI24/00192 Grant and Generalitat de Catalunya (2021 SGR 00776).

The funding sources had no role in the design and conduct of the study, interpretation of the data, writing of the manuscript, and decision to submit the manuscript for publication.

## Author contributions

The study was designed by A.L.A., D.A.E., E.D.P., K.P., and P.Č. E.D.P., and P.Č. conceptualized and supervised the project. A.Č., B.H., D.G., E.H., K.J., S.A.C., S.Sh., S.Sz., S.Sm., S.G., and T.O. sectioned FFPE tissues. B.H., D.G., E.H., H.O., I.V., K.J., S.Sm., S.G., S.H., and T.O. conducted histological staining. B.H., D.G., E.H., K.J., S.Sm., S.G., and T.O. performed laser capture microdissection. B.H., D.G., E.H., K.J., M.K., M.R., S.Sm., S.G., and T.O. annotated H&E images. D.A.E. provided pathologist review of cases, including annotations and TILs assessment. A.Č., A.S., B.H., D.G., E.H., H.O., I.V., K.J., S.Sz., S.Sm., S.G., S.H., T.O., and Z.F. conducted RNA-FISH. A.Č., A.S., B.H., D.G., E.H., K.J., S.Sz., S.Sm., S.G., and T.O. quantified RNA-FISH images. E.H., I.V., L.B., M.K., M.R., and S.Sm. extracted RNA and conducted RNA quality control. L.B., M.K., M.R., N.M., and S.H. prepared RNA-Seq libraries and conducted cDNA quality control. L.B., M.K., M.R., and N.M. pooled libraries for sequencing. M.G. and F.N. designed and developed predictive and prognostic gene expression signatures, including model development and validation. E.D.P., F.N., M.G., and S.G. conducted bioinformatic and statistical analyses. B.H., D.G., D.A.E., E.D.P., F.N., K.J., L.B., M.K., M.R., M.G., N.M., S.Sm., S.G., T.O., and P.Č. verified the underlying data, analyzed, and interpreted the data, prepared figures, and wrote the manuscript. B.H., D.A.E., E.D.P., G.R., K.J., K.Je., L.B., M.G., S.A.C., S.H., T.O., and P.Č revised the manuscript. A.L.A., F.C., F.L., G.E., G.R., I.A., I.S., J.A., K.Je., K.P., K.T., L.C., M.H., P.G., P.R., R.N., R.R., S.Sh., and S.Se. acquired the samples. E.D.P., B.H., D.G., A.L.A., F.C., F.L., G.E., G.R., I.A., I.S., J.A., K.Je., K.P., K.T., L.C., M.H., P.G., P.R., R.N., R.R., S.Sh., and S.Se. curated clinical data. All authors had full access to the data, provided critical comments and feedback on the manuscript, and accepted responsibility to submit the manuscript for publication.

## Ethics & Inclusions

This study complies with all relevant ethical regulations for human research participants in accordance with the Declaration of Helsinki. The collection and use of human tissue and patient data in this real-world, multicenter, retrospective cohort of 590 patients diagnosed with eTNBC and treated with neoadjuvant chemotherapy with or without immunotherapy was approved by the Ethics Committee of the Bratislava Self-Governing Region (Ref. No. 05320/2020/HF). In addition, the study protocol was approved by local ethical committees for each of the participating sites that provided anonymized patient specimens and clinical data. Each site obtained informed consent from the patients for inclusion in the study. No discrimination occurred in the selection of patients and both clinical partners and MultiplexDX ensured sensitive patient information was anonymized when appropriate, secured both physically and electronically, and was processed in accordance with GDPR.

---

**Extended Data Figure 1.**
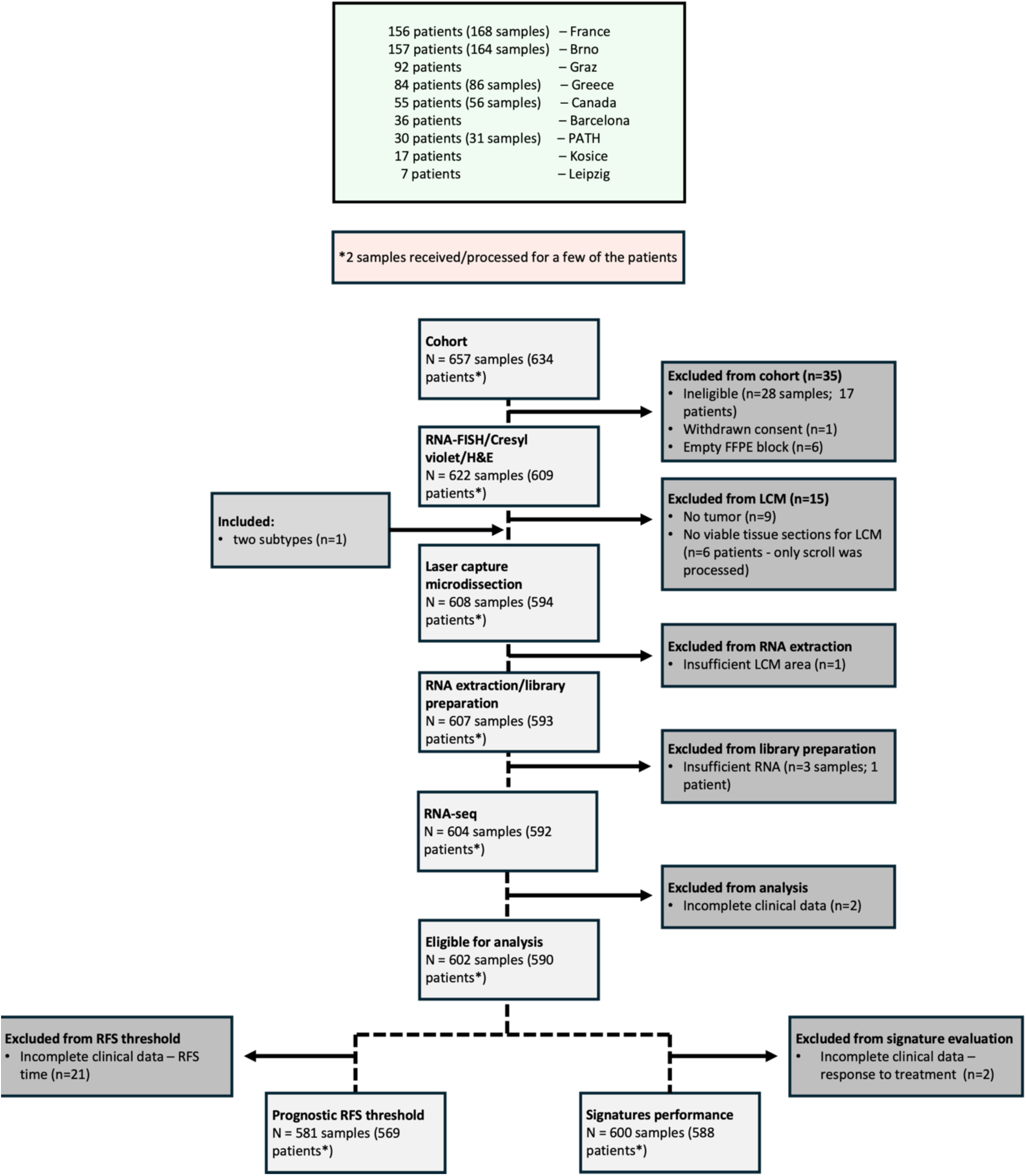
A CONSORT flow diagram depicting the number of diagnostic core biopsy samples and unique patients at each stage of the study. Samples excluded from the study are broken down into pre-analytical and analytical factors. Preanalytical factors are related to issues from the specimens and the site of origin such as ineligibility due to exclusion/inclusion criteria, withdrawn informed consent, specimens identified as not having any or insufficient tissue or tumor in the FFPE block, and missing clinical data that prevent the specimens from being used in downstream analyses. Analytical factors include issues pertinent to the Multiplex8+ wet- and dry-laboratory workflow, including insufficient RNA from extraction, poor quality sequencing data, and statistical outliers.

**Extended Data Figure 2.**
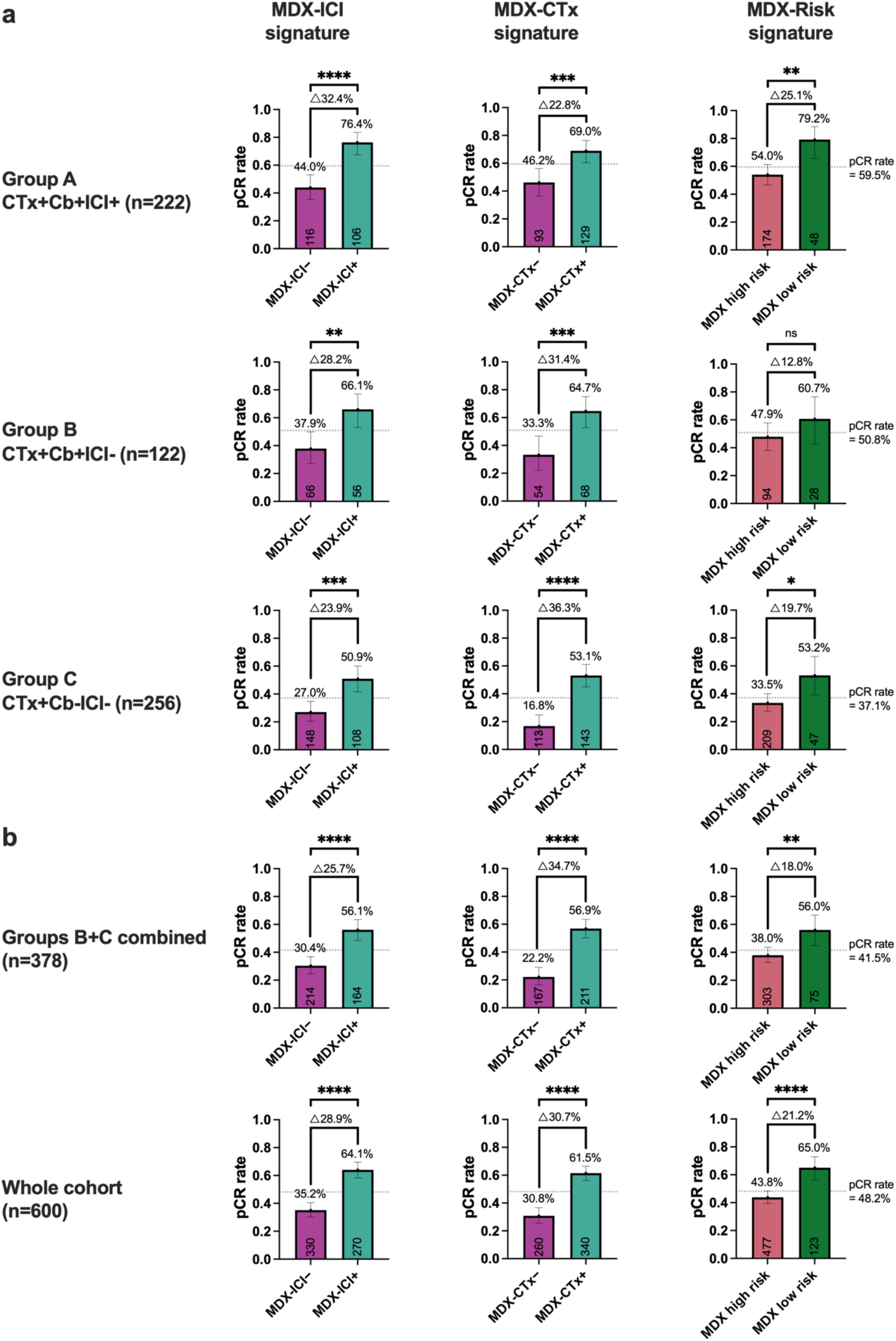
Bar charts show the pCR rates for each of the three MDX signatures in the three treatment groups, the combined CTx+Cb+ICI− and CTx+Cb−ICI− group, and the overall cohort. **a**) The pCR rates for patients that are either negative or positive for the MDX-ICI, MDX-CTx, and MDX-Risk signatures in the CTx+Cb+ICI+ group, CTx+Cb+ICI− group, and the CTx+Cb−ICI−. **b**) The pCR rates for the same groups in panel **a**) in the combined CTx+Cb+ICI− and CTx+Cb−ICI− group and the overall cohort. The dotted lines denote the overall pCR rate of the group irrespective of biomarker subgroup. Text above each bar shows the pCR rates for patients classified as either biomarker – or +. The connecting lines show the difference (delta Δ) in pCR rates between biomarker – or + groups. The numbers within each bar near the x-axis show the number of patients in each biomarker class. The error bars show the 95% confidence intervals (CI).

**Extended Data Figure 3.**
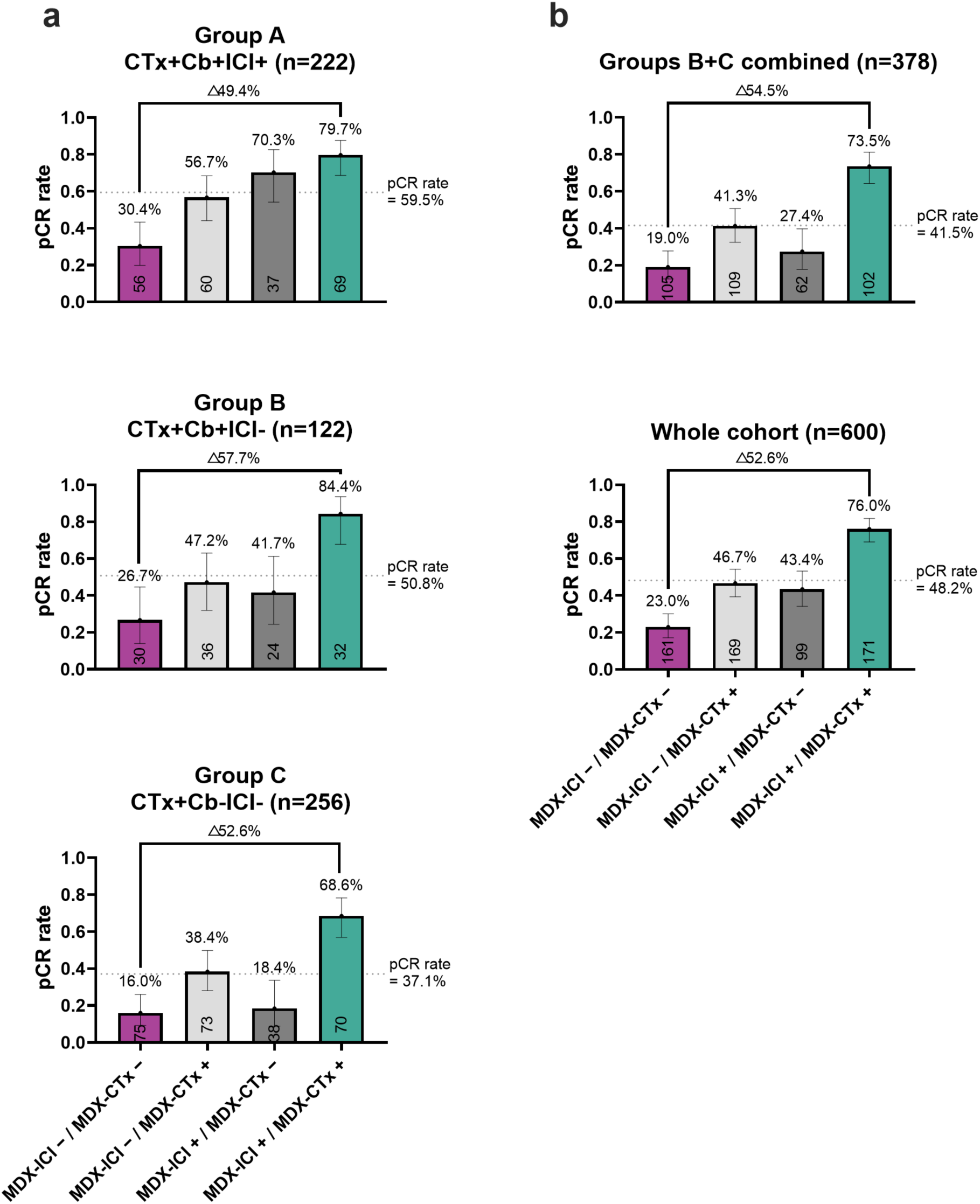
Bar charts show the pCR rates after combing the MDX-ICI and MDX-CTx pCR response signatures, yielding four subgroups: ICI + / CTx + (positive for both signatures), ICI – / CTx + (negative for MDX-ICI but positive for MDX-CTx), ICI + / CTx – (positive for MDX-ICI but negative for MDX-CTx), and ICI – / CTx – (negative for both signatures). **a**) The pCR rates in the CTx+Cb+ICI+ group, CTx+Cb+ICI− group and CTx+Cb−ICI−. **b**) The pCR rates in the combined CTx+Cb+ICI− and CTx+Cb−ICI− group and the overall cohort. The dotted lines denote the overall pCR rate of the arm irrespective of biomarker subgroup. Text above each bar shows the pCR rates for each group. The connecting lines show the difference (delta Δ) in pCR rates between ICI + / CTx + or ICI – / C – groups. The numbers within each bar near the x-axis show the number of patients in each biomarker class. The error bars show the 95% confidence intervals (CI).

**Extended Data Figure 4.**
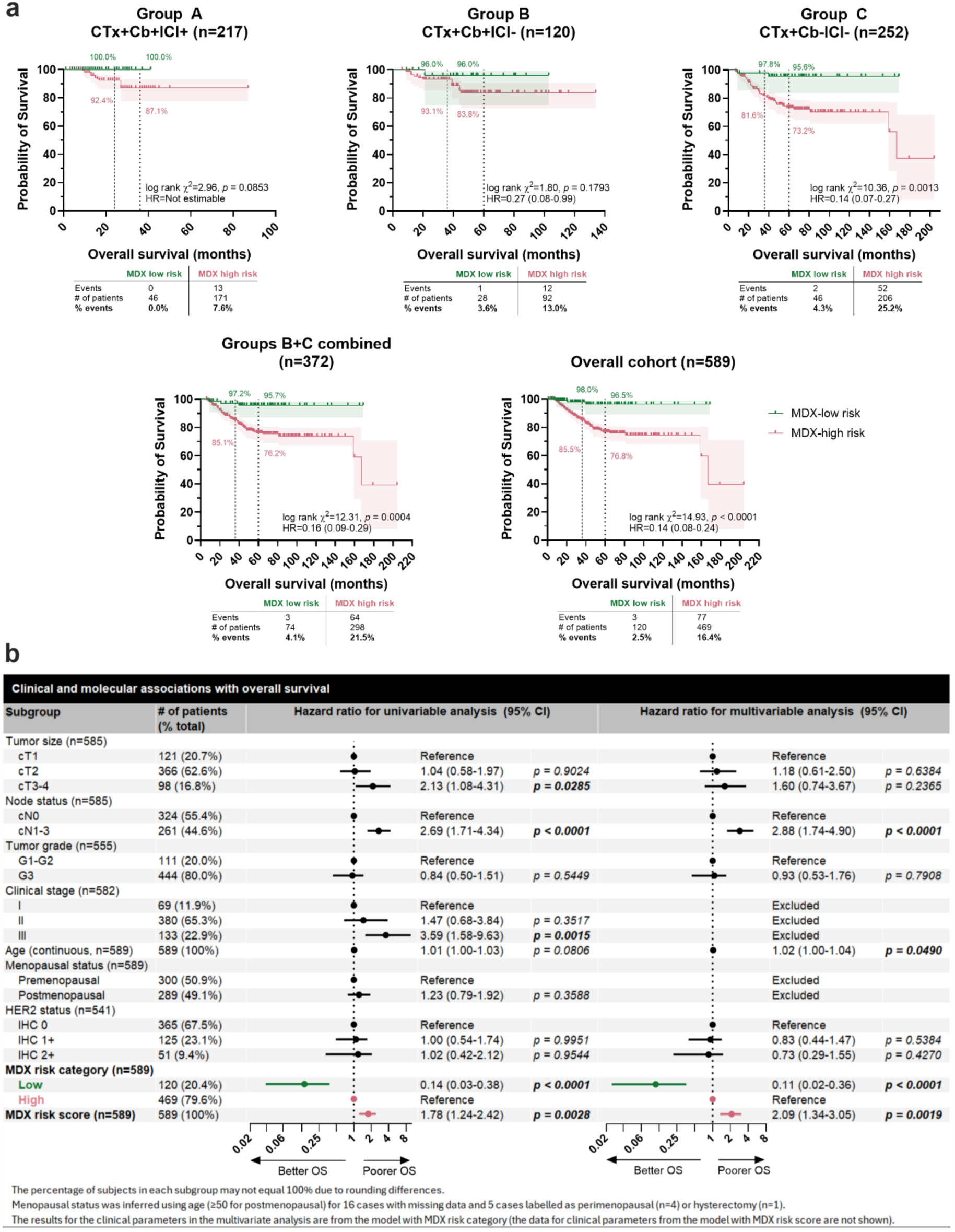
The MDX prognostic signature associates with overall survival (OS) in early TNBC. **a**) Kaplan-Meier curves show the probability of OS in patients classified as either MDX high or low risk assessed in the CTx+Cb+ICI+ group, CTx+Cb+ICI− group, CTx+Cb−ICI− group, combined CTx+Cb+ICI− and CTx+Cb−ICI− group, and the overall cohort. The dotted lines denote 2-yr and 3-yr OS for the CTx+Cb+ICI+ group and 3-yr and 5-yr OS for all other groups. The colored annotations show the probability of OS for each risk group at the respective time points. The shaded areas show the 95% confidence interval (CI) of each curve. A logrank (Mantel-Cox) test was used to assess statistical differences between curves and the logrank method was used to derive hazard ratios. The table below each Kaplan-Meier curve shows the number of events (deaths), total patients (censored+events), and the percentage of events. **b**) Univariable and multivariable Cox proportional hazards models illustrate the relationship between key clinical parameters and the MDX prognostic signature score (a continuous variable) and the corresponding MDX high and low risk groups with OS. Forest plots show the hazard ratio point estimates and 95% confidence interval (CI). Statistically significant (*p* < 0.05) comparisons are in bold text.

**Extended Data Figure 5.**
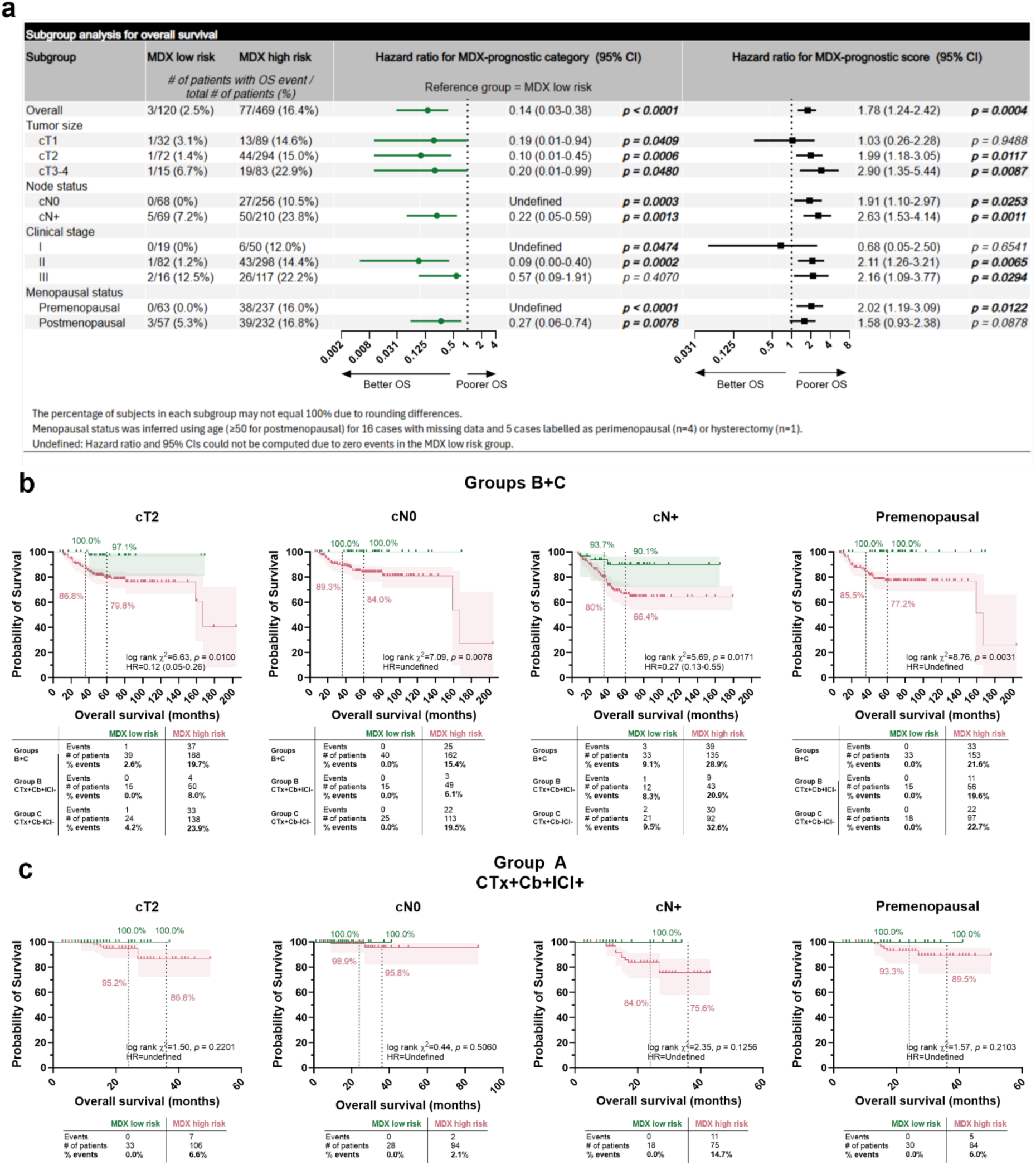
The MDX prognostic signature is strongly associated with overall survival (OS) in key clinical subgroups. Univariable Cox proportional hazards models illustrate the relationship between the MDX high and low risk groups and corresponding MDX prognostic signature score (a continuous variable) with OS in a subgroup analysis. Forest plots show the hazard ratio point estimates and 95% confidence interval (CI). Statistically significant (*p* < 0.05) comparisons are in bold text. **b-c**) Kaplan-Meier curves show the probability of OS in patients classified as either MDX high or low risk assessed in the combined CTx+Cb+ICI− and CTx+Cb−ICI− group (**b**) and CTx+Cb+ICI+ group (**c**) for exemplary clinical subgroups. The dotted lines denote 3-yr and 5-yr OS for the combined CTx+Cb+ICI− and CTx+Cb−ICI− group and 2-yr and 3-yr OS for the CTx+Cb+ICI+ group. The colored annotations show the probability of OS for each risk group at the respective time points. The shaded areas show the 95% confidence interval (CI) of each curve. A logrank (Mantel-Cox) was used to assess statistical differences between curves and the logrank method was used to derive hazard ratios. The table below each Kaplan-Meier curve shows the number of events (deaths), total patients (censored+events), and the percentage of events.

**Extended Data Figure 6.**
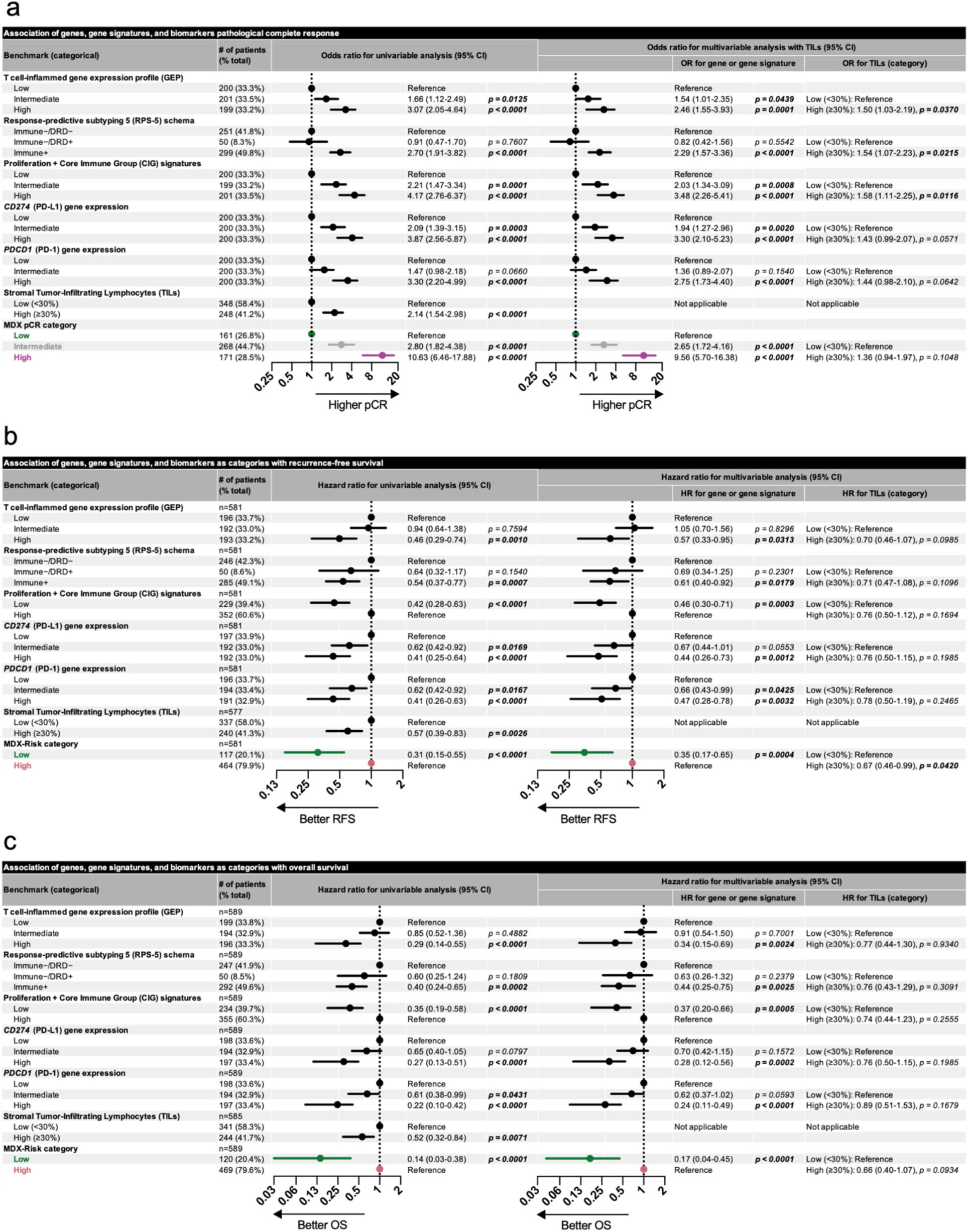
Benchmarking MDX pCR probability groups and MDX-Risk groups against other molecular biomarkers. **a-c**) Association of various molecular biomarkers with pCR (**a**), RFS (**b**), and OS (**c**) in both univariable and multivariable models. Multivariable models contained TILs as a categorical variable in addition to each benchmark ran in separate models. Forest plots show either the odds ratio (pCR) or hazard ratio (RFS/OS) point estimates and 95% confidence intervals (CI) of the various molecular biomarkers. Statistically significant (*p* < 0.05) comparisons are in bold text.

**Extended Data Figure 7.**
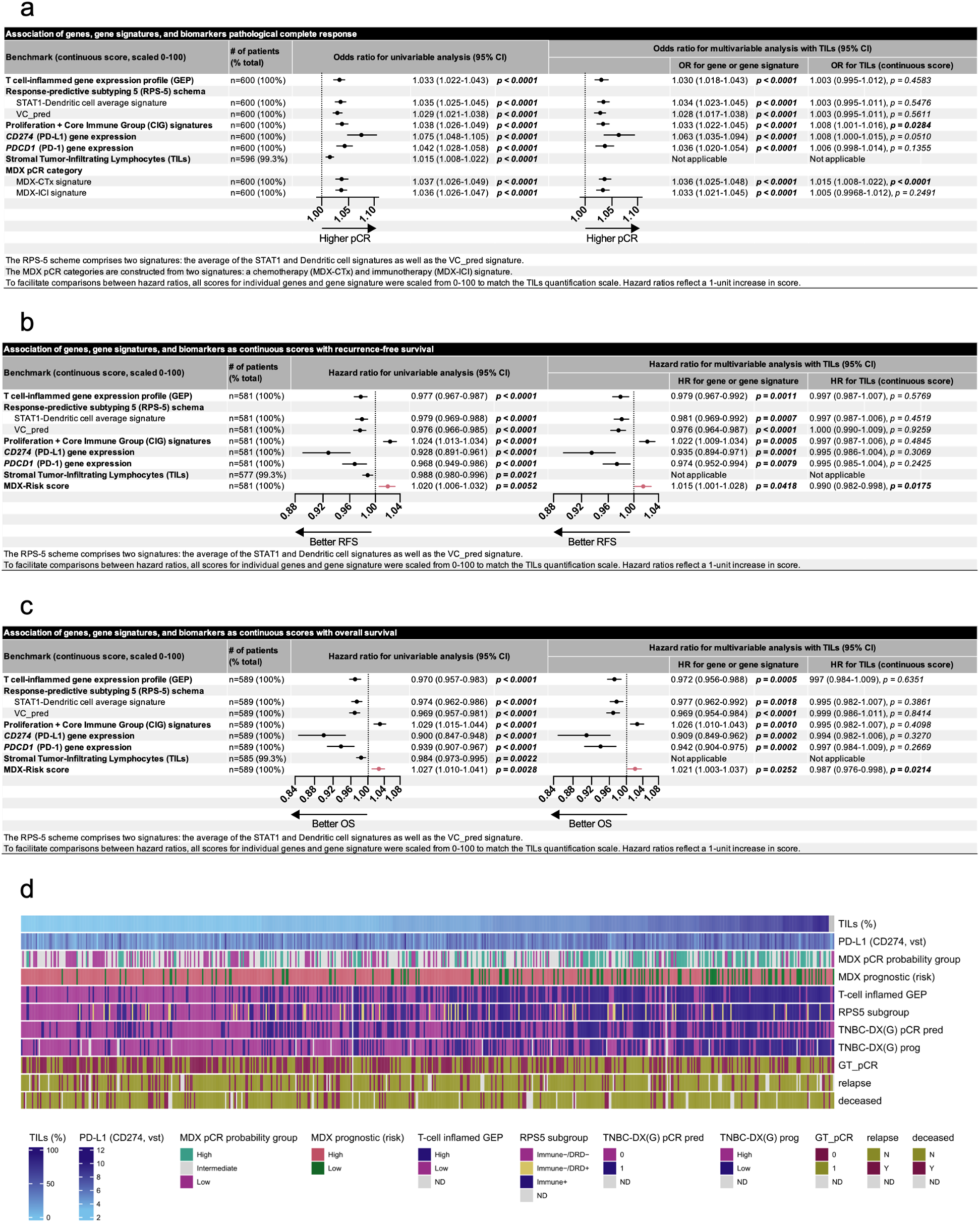
Benchmarking MDX-CTx and MDX-ICI signatures and the MDX-Risk signature against other molecular biomarkers all expressed as continuous scores. **a-c**) Association of molecular biomarkers with pCR (**a**), RFS (**b**), and OS (**c**) in both univariable and multivariable models. Multivariable models contained TILs as a continuous score in addition to each benchmark ran in separate models. Forest plots show either the odds ratio (pCR) or hazard ratio (RFS/OS) point estimates and 95% confidence intervals (CI) of the various molecular biomarkers. Statistically significant (*p* < 0.05) comparisons are in bold text. All continuous scores were min-max scaled from 0-100 to ensure hazard ratios reflect a standardized metric. **d**) Heatmap shows the classifications of each benchmark with neoadjuvant response and long-term outcomes. Samples are sorted by increasing TILs percentage.

**Extended Data Figure 8.**
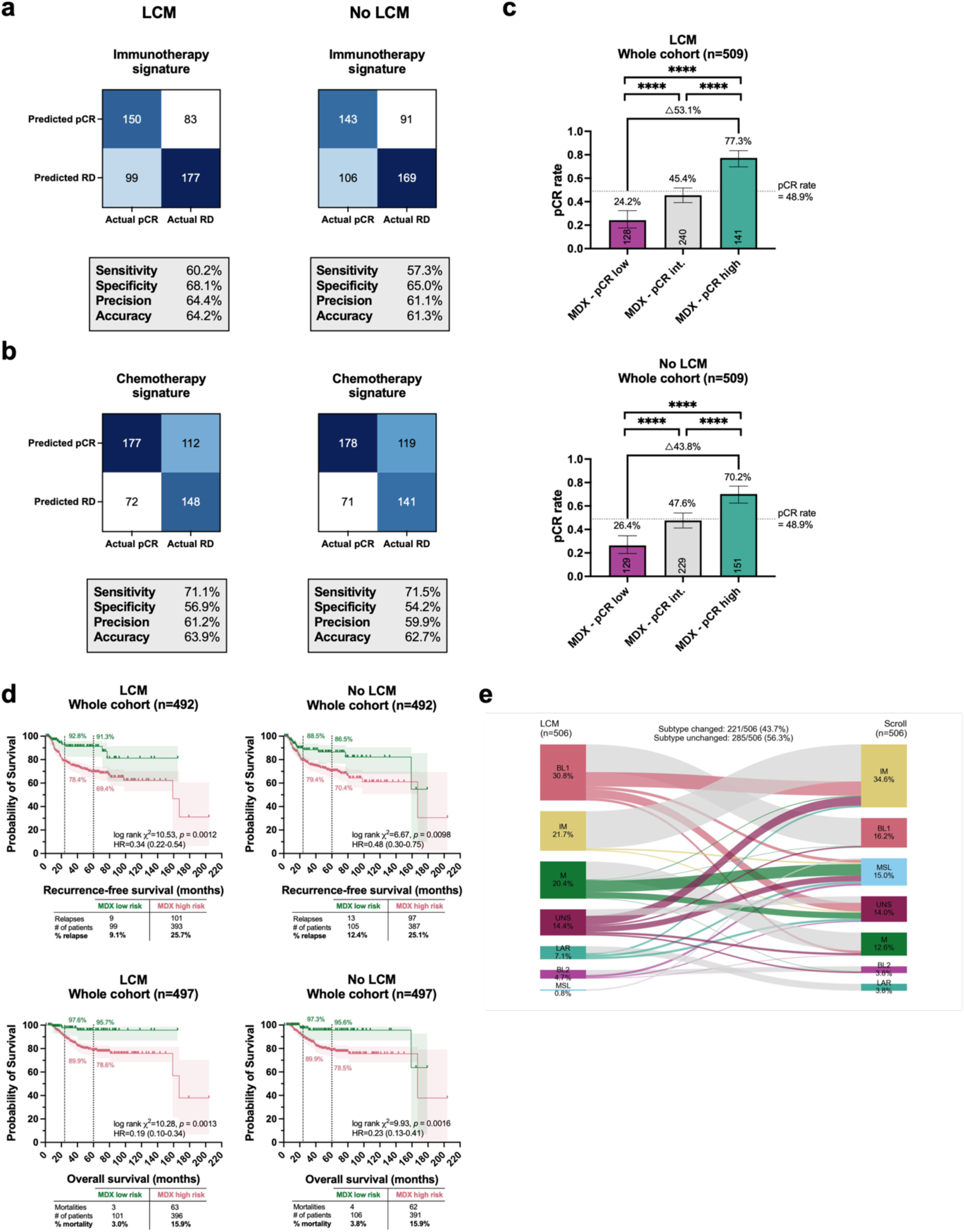
Laser capture microdissection (LCM) improves stratification of patients according to clinical outcomes. **a-b**) Confusion matrices show the performance of the MDX-ICI (**a**) and MDX-CTx (**b**) signatures in paired samples that were either laser capture microdissected (LCM) or processed as a bulk-specimen (No LCM). **c**) Bar charts show pCR rates in the MDX pCR low, intermediate, and high probability groups in LCM vs no LCM samples. **d**) Kaplan-Meier curves compare LCM vs no LCM in stratifying patients into MDX-Risk low- and high-risk groups for RFS (top row) and OS (bottom row). The dotted lines denote 2-yr and 5-yr RFS/OS and colored annotations show the probability of RFS/OS for each group at the respective time points. The shaded areas show the 95% confidence interval (CI) of each curve. A logrank (Mantel-Cox) test was used to assess statistical differences between curves and the logrank method was used to derive hazard ratios. The table below each Kaplan-Meier curve shows the number of events (recurrence events or deaths), total patients (censored+events), and the percentage of events. (**e**) The Sankey diagram shows the distribution of TNBC subtypes in paired samples that underwent LCM or No LCM.

