## SUPPLEMENTARY INFORMATION for "Spatially informed comprehensive tumor transcriptomic profiling stratifies clinical outcomes in early triple negative breast cancer"

### Section I. Development of Multiplex8+ signatures to stratify clinical outcomes in patients with eTNBC

*Development of the Multiplex8+ chemotherapy response signature*

For the chemotherapy (CTx) response signature (MDX-CTx), we deployed a pipeline published by Atreya and colleagues (**Supplementary Fig. 1**)[1] on a harmonized compendium of microarray gene expression datasets (GSE205568) from 23 studies involving 706 patients diagnosed with eTNBC and who received neoadjuvant anthracycline/taxane-containing regimens[2]. We specifically used the quantile normalized pooled dataset with a total of 9,184 genes present in all datasets. In the first section of the pipeline, we selected a list of stable features, with a few changes from the published pipeline. We used Synthetic Minority Over-sampling Technique (SMOTE) to balance the classes[3] and then performed a Mann-Whitney U test to statistically compare gene expression distributions between samples that had a pathological complete response (pCR) versus those with residual disease (RD). Only genes with a relaxed cutoff (*p-value* < 0.01) were retained in a pooled feature list for recursive feature elimination (RFE).

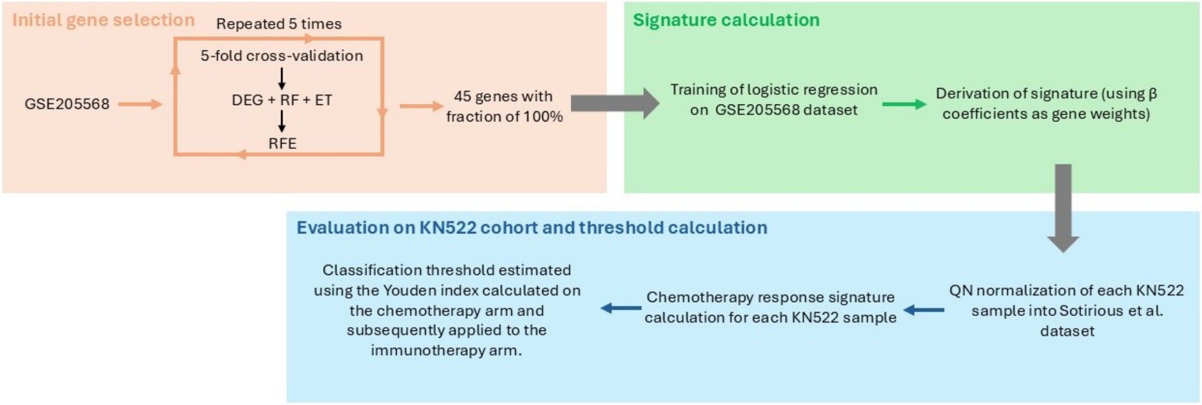


**Supplementary Figure 1. Schematic overview of the key steps involved in chemotherapy response signature development.** The approach consists of three stages: 1. Initial gene selection using a previously published machine learning pipeline; 2. Derivation of a logistic regression signature; and 3. Evaluation of the final gene signature (genes and algorithm weights) on the real-world, retrospective cohort (e.g., KN522 cohort) of 590 patients (602 samples) diagnosed with early triple-negative breast cancer (eTNBC). DEG, differentially expressed genes; RF, random forest; ET, ExtraTreeClassifier; RFE, recursive feature elimination.

For RFE-based feature selection, we used Random Forest[4] and the ExtraTreeClassifier[5] and the results of the genes selected within folds confirm their predictive signal with Area under the Receiver Operating Characteristic (AUC) curves ranging from 0.73-0.74 (**Supplementary Fig. 2**). A list of all 2,404 genes that passed the initial gene selection filter at least once, together with their fraction scores, are reported in **Supplementary Data 1**.

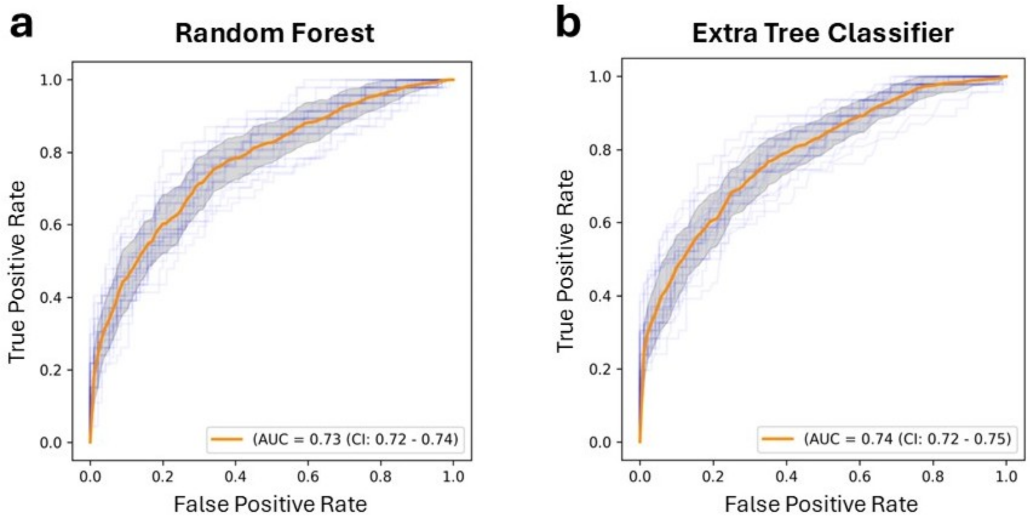


**Supplementary Figure 2. Area under the Receiver Operating Characteristic (AUC) curves of models used for RFE-based feature selection in chemotherapy response signature development. a,** Random Forest and **b,** Extra tree classifier used for initial gene selection in the development of the MDX-CTx signature. ROC curves from the genes selected in each of the 25 folds of repeated stratified cross-validation (CV) are shown in light blue. The orange curve represents the mean ROC curve of each fold with 95% CIs shown in the shaded gray regions.

In the iterative feature selection step, we only retained genes selected across all folds and iterations of the pipeline (i.e., genes having a feature fraction of 1), which yielded 45 robust genes. Subsequently, in the second part of the pipeline, we trained the final logistic regression model and derived the respective gene signature weights using and the same training dataset (GSE205568) as for the gene selection step[2]. For validation, to overcome the mismatch between the scale of expressions of the training and validation datasets, we quantile normalized the TPM gene expression data to the reference (training) dataset. This was followed by *z-score* normalization using the mean and standard deviation for each gene calculated from the training dataset. We used the argument class_weight="balanced" to account for class imbalance in the training dataset. The gene weights were derived from the beta coefficient of the logistic regression, with the final signature expressed as:

$$Signature_i = \sum_{g \epsilon G} X_{i,g} \times \beta_g$$

where:

$Signature_i$ is the continuous signature score for sample *i*

$X_{i,g}$ is the expression value of gene *g* in sample *i*

$\beta_g$ is the weight (logistic regression coefficient) for gene *g*

For finding the optimal signature threshold on the validation dataset, we used Youden's index (*J*) that balances sensitivity and specificity[6]. The threshold was set based on the highest Youden's index in eTNBC patients that were treated with chemotherapy (i.e., CTx+Cb+ICI− and CTx+Cb−ICI− groups B+C) and subsequently applied on to patients in the immunotherapy group A (CTx+Cb+ICI+). The results from the validation are presented in the main text.

*Development of the Multiplex8+ immunotherapy response signature*

To develop an immunotherapy (ICI) response signature (MDX-ICI), we leveraged a large meta-analysis of transcriptomic data from 16 cohorts with a total of 1,739 eTNBC samples that identified genes associated with neoadjuvant response and long-term outcomes[7]. This initial pool consisted of 1,942 high-quality genes that were reported to be significantly associated with either pCR/RD or favorable/unfavorable disease-free survival (i.e., all four quadrants in the meta-analysis) genes after integration of TNBC NAC and DFS meta-analysis. Next, we verified the presence of these genes in our training datasets, which consisted of the pembrolizumab (n=29; GSE194040[8]) and the durvalumab plus olaparib (n=21; GSE173839[9]) arms of the phase II, adaptive I-SPY2 platform trial as well as our real-world, retrospective study (n=590 patients, n=602 samples) validation dataset, yielding 1,742 genes present in both the training and validation datasets that were retained for model development (**Supplementary Fig. 3**).

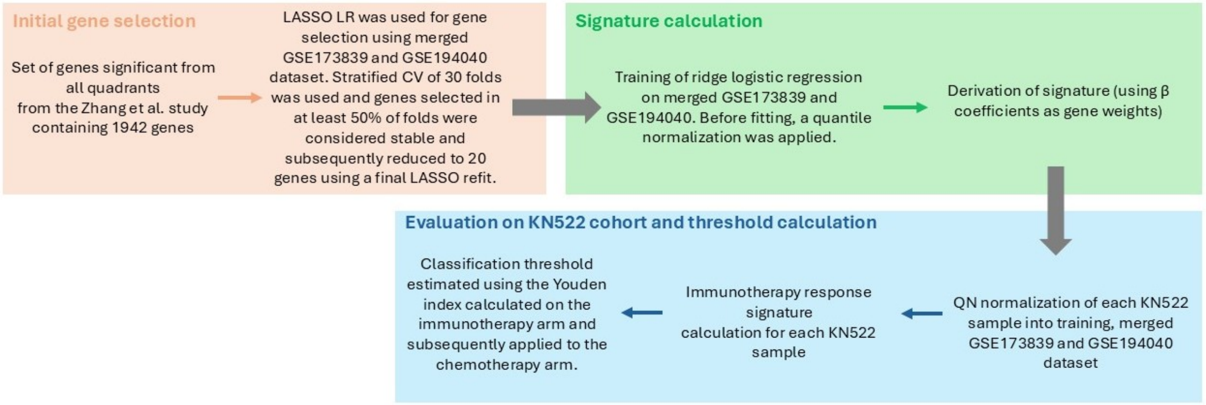


**Supplementary Figure 3. Schematic overview of the key steps involved in the training and validation of the MDX-ICI signature.** The approach consists of three stages: 1. Initial gene selection from a TNBC transcriptomic meta-analysis followed by a global regularized (LASSO) logistic regression to refine the list; 2. Training of a ridge (L2 regularized) logistic regression signature and derivation of gene weights; and 3. Evaluation of the final gene signature (genes and algorithm weights) on the real-world, retrospective cohort (e.g., KN522 cohort) of 590 patients (602 samples) diagnosed with early triple-negative breast cancer (eTNBC). CV, cross-validation; LR, logistic regression; QN, quantile.

For initial feature selection, we used a global penalized (LASSO) logistic regression on the 1,742 genes present in the training and validation sets.[10]. Genes were z-score standardized and weights were adjusted due to imbalanced classes as described for the MDX-CTx signature. For gene selection as well as training, we used the merged pembrolizumab (n=29; GSE194040[8]) and durvalumab plus olaparib (n=21; GSE173839[9]) arms of I-SPY2. LASSO regularization strength was tuned by inner 3-fold stratified cross-validation (CV), using the Area under the Receiver Operating Characteristic AUROC curves as the selection criterion (**Supplementary Fig. 4**) and gene stability was calculated using repeated stratified CV with 3 folds repeated 10 times (**Supplementary Data 2**). Genes with non-zero coefficients were counted in each outer fold, and genes selected in at least 50% of the folds were considered stable. This approach produced 22 stable genes. A final LASSO refit on the full training cohort reduced this to 20 final genes.

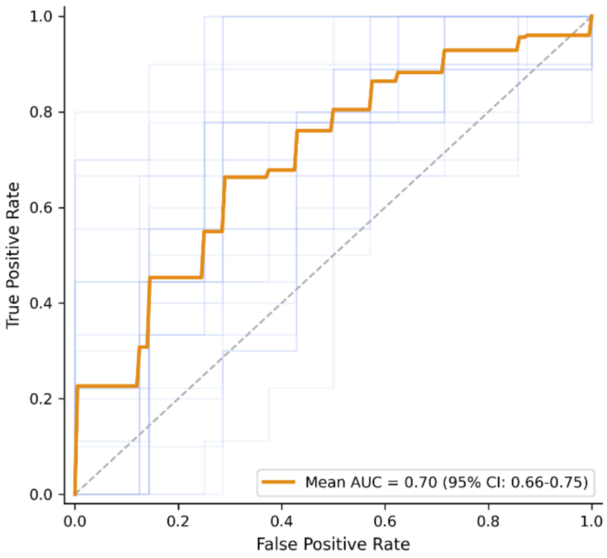


**Supplementary Figure 4. Receiver Operating Characteristic (ROC) curve during LASSO feature selection.** ROC curves from 30 folds of repeated stratified cross-validation (CV) are shown in light blue. The orange curve represents the mean ROC curve with mean AUC 0.70 and 95% CI of 0.66-0.75.

For training and derivation of the final signature weights, we used logistic regression applied on quantile-normalized and standardized data. Before model training, only the 20 genes selected from LASSO regression were quantile normalized using all samples in the merged training cohort. The final ridge regression model was optimized across 3-fold CV. Finally, the continuous MDX-ICI signature score is the logistic regression decision function (like the MDX-CTx signature) and the threshold was estimated using Youden's index in the real-world, retrospective (n=590 patients, n=602 samples) validation cohort. Here, the threshold was determined by the highest Youden's index in eTNBC patients that were treated in the immunotherapy group A (CTx+Cb+ICI+) and subsequently applied to patients in the chemotherapy (i.e.,

CTx+Cb+ICI− and CTx+Cb−ICI−) groups B+C). The results from the validation are presented in the main text.

*Development of the Multiplex8+ prognostic signature*

The Multiplex8+ prognostic signature (MDX-Risk) was developed using the stable Cox regression approach[11]. The method consists of an independently operated sample weighting module and a sample weighted Cox regression model (**Supplementary Fig. 5**).

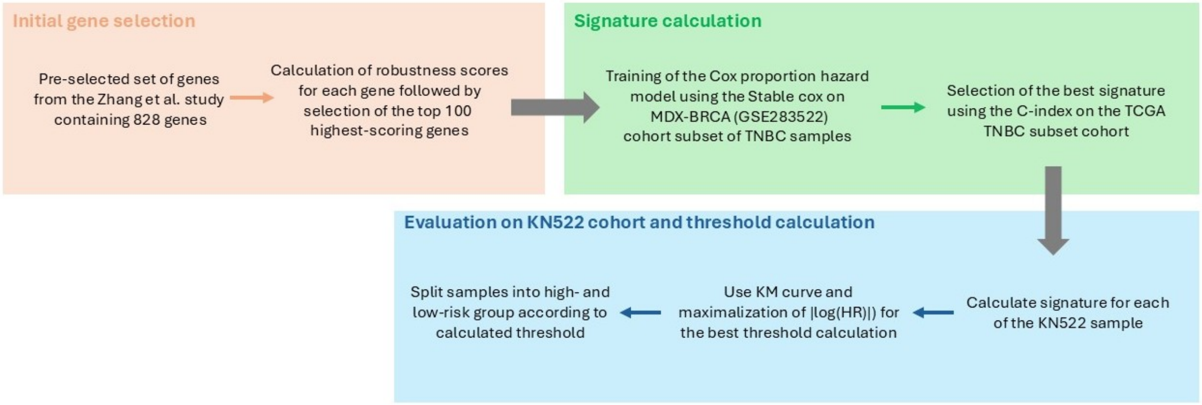


**Supplementary Figure 5. Schematic overview of the key steps involved in the development of the MDX-Risk signature.** The workflow consists of three stages: 1. Initial gene selection from a TNBC transcriptomic meta-analysis, calculation of robustness scores, followed by selection of the top 100 scoring genes; 2. Training of the Cox proportional hazards model and model selection based on C-index; and 3. Evaluation of the final gene signature (genes and algorithm weights) on the real-world, retrospective cohort (e.g., KN522 cohort) of 590 patients (602 samples) diagnosed with early triple-negative breast cancer (eTNBC). KM, Kaplan-Meier; HR, Hazard ratio.

The advantage of this method over classical Cox regression is the use of an independent sample weighting algorithm with the ability to weight samples in a way that removes spurious feature correlations, while providing theoretical guarantees for identifying stable features with generalization ability across independent cohorts. For sample weighting, we used a method described in the original paper, the Sample Reweighted Decorrelation Operator (SRDO)[12].

For initial gene selection, we used the transcriptome meta-analysis of breast cancer response to neoadjuvant therapy[7] like we for the MDX-ICI signature. However, we focused on a specific set of 828 genes associated with favorable clinical outcomes (increased pCR and favorable DFS, quadrant 1). We reduced the 828 genes to a subset of 100 genes using the following criteria: $-log_{10}(adjusted\ p\ value)$; effect size; ratio of coherent studies to total studies. For each gene, we summarized these criteria to the gene's RobustnessScore as the sum of these three features. This score quantifies the overall confidence of the gene's association with increased pCR and favorable DFS. Subsequently, genes were ranked in descending order, with genes with the highest scores being those with the strongest and most consistent positive effect, and we selected the top 100 genes with the highest RobustnessScore (see **Supplementary Data 3**).

For training and validating (step 2), we used two large international cohorts of patients with eTNBC treated in the adjuvant setting, including the MDX-BRCA[13] (training set, n=159) and TCGA-BRCA[14,15] (validation set, n=148) cohorts. The long-term endpoints were progression-free survival and DFS for MDX-BRCA and TCGA-BRCA, respectively. The first step of our stable cox pipeline is an initial quality check of the genes. Only genes with an average TPM value of > 1 and a rate of zero expression < 0.5 (i.e., <50% of samples have zero expression based on raw counts) passed the initial filter before training. After quality control, 96 genes were retained, normalized using *z-scores* based on the training dataset, with the mean and standard deviation of the training dataset applied to the validation (TCGA-BRCA) and test (real-world, retrospective eTNBC) datasets. We decided to omit the initial univariate Cox model described in the original method, as we already pre-selected 100 genes making it unnecessary. The samples were reweighted using all the genes that passed the quality control, followed by a weighted Cox. The weighted Cox absolute beta coefficient is then used to rank the genes in descending order (**Supplementary Data 4**). These are then fed into a standard (unweighted) Cox model to find the optimal number of genes for the signature. We tuned the lambda coefficients of weighted cox as well as the final unweighted cox. Moreover, we also tuned the SRDO hidden layer sizes and clipping bounds for sample weights after SRDO. The models are continuously tuned based on the C-index of the independent TCGA validation cohort (**Supplementary Fig. 6**). After finding the optimal number of genes and hyperparameters, the final standard (unweighted) Cox is trained.

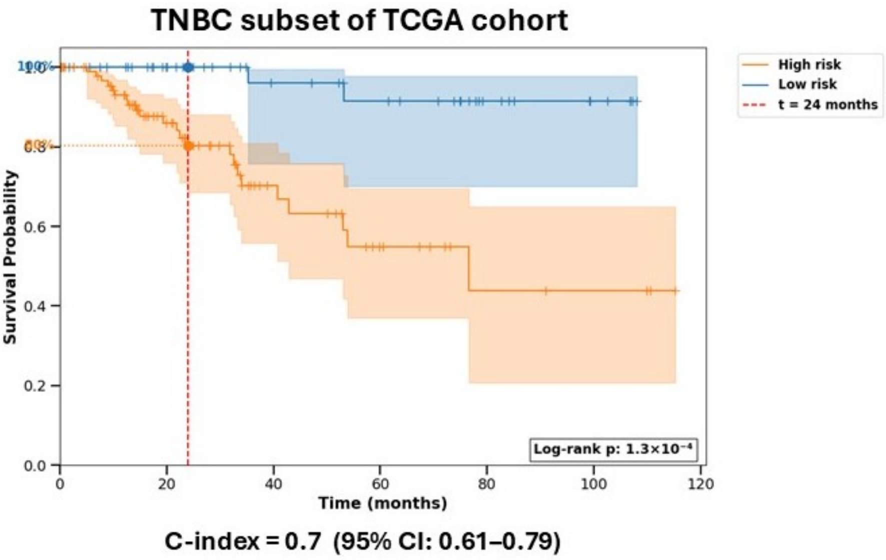


**Supplementary Figure 6. Association of the MDX-Risk signature with disease-free survival (DFS) in the TCGA-BRCA cohort.** Patients are stratified into high- and low-risk groups using the MDX-Risk signature by using a threshold that maximizes |log(HR)|). Red dashed line represents patient DFS probability after 2 years. The colored annotations show the probability of RFS for each risk group at 2 years. The shaded areas show the 95% confidence interval (CI) of each curve.

The beta coefficients of the genes from the standard model are then used to calculate the score of the signature itself. The prognostic signature can be expressed as:

$$Signature_i = exp\left(\sum_{g \in G} X_{i,g} \times \beta_g\right)$$

where:

$X_{i,g}$ is the expression value of gene *g* in sample $i$

$\beta_g$ is the weight (coefficient) of gene $g$

$G$ is the set of genes

The signature score represents the relative risk for sample *i* compared to a baseline of 1.

Classification of patients into high- and low-risk groups was performed using a threshold of the MDX-Risk signature score that was optimized using a method that maximized the absolute value of the log hazard ratio (|log(HR)|). This corresponds to the largest relative difference in hazard between the high- and low-risk group. The optimal threshold was determined by scanning 500 candidate cutoffs from the 10th–90th percentiles of the signature score distribution. For each potential threshold, samples were stratified into high- (score ≥ threshold) and low-risk (score < threshold) groups. Only a threshold with at least 20% of the total cohort samples was considered valid to avoid a one-sided stratification. For every threshold, we fitted a Cox model with the binary group variable and calculated the log hazard ratio. The optimal threshold was used to stratify patients via Kaplan-Meier plots and risk group differences were analyzed using the log-rank *p*-value and C-index[16]. The optimal threshold was finetuned and test on the real-world, retrospective (n=590 patients, n=602 samples) test cohort independently using both recurrence free survival and overall survival as endpoints. The results from the validation are presented in the main text.

### Section II. Response and Prognosis Profile: A hypothesis generating framework for identifying patients for treatment optimization paradigms.

*Stratification of MDX pCR probability groups with the MDX-Risk groups*

Despite being trained on different cohorts and clinical endpoints, as well as containing minimal overlap in genes (**Supplementary Fig. 7a**), there is noticeable overlap on the patient subgroups identified by the MDX-Risk and MDX pCR probability groups. For example, MDX low and intermediate pCR groups are enriched in patients with MDX-high risk and have higher MDX-Risk signature scores (**Supplementary Fig. 7b-c**). The MDX-Risk groups have different pCR rates (**Extended Data Fig. 2**), while the MDX pCR probability groups have different RFS and OS (**Supplementary Fig. 7d**).

Given the composition of high and low risk patients in the MDX intermediate and high pCR groups, we investigated whether the MDX-Risk signature could further stratify these groups. The high and low risk MDX intermediate group had strikingly different outcomes, with patients classified as MDX-low risk having better prognoses in both RFS and OS as well as a 18.1% difference in pCR rates (**Supplementary Fig. 7e**). In the MDX high pCR group, the MDX-Risk signature showed a modest contribution to outcomes (**Supplementary Fig. 7f**).

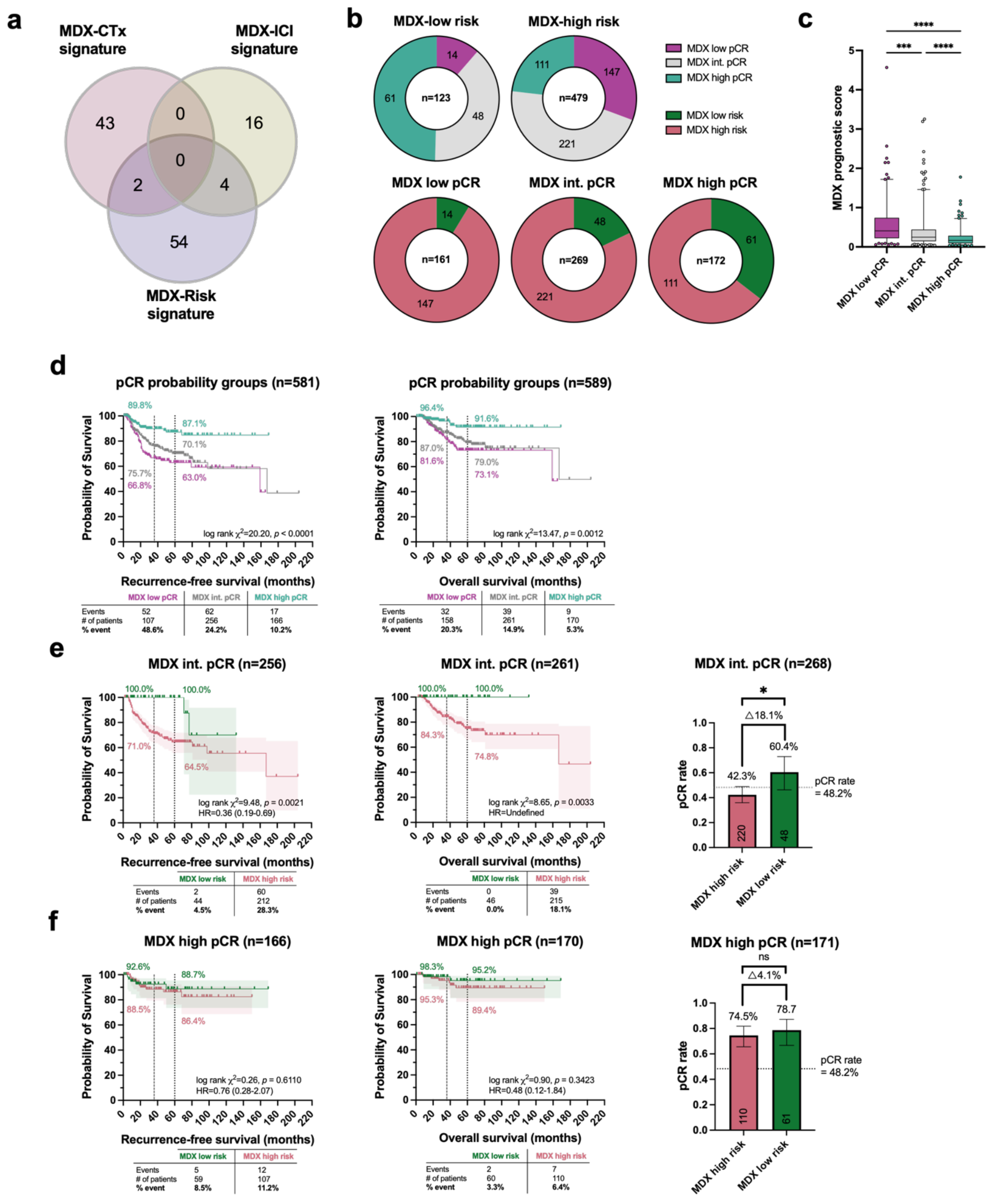


**Supplementary Figure 7.** MDX pCR probability and prognostic groups are associated and synergize to provide more granular stratification. **a**) Overlap of genes in the MDX chemotherapy, immunotherapy, and prognostic signatures. **b**) Proportion of patients classified as low, intermediate, and high pCR probability in either low- or high-risk groups (top figures) as well as proportion of patients classified as low or high risk in either low, intermediate, or high pCR probability groups (bottom figures). **c**) The MDX prognostic signature scores in each of the three pCR probability groups. Boxes show the interquartile range, dotted line in each box denotes the median, whiskers show the 95% confidence intervals, and dots show outliers. One-way ANOVA with Tukey's post hoc multiple comparisons test, ****$p$ <0.0001. **d-f**) Kaplan-Meier curves show the probability of RFS and

OS in patients classified as either MDX low, intermediate, or high pCR probability in the overall cohort (**d**) as well as the MDX intermediate (**e**) and high (**f**) pCR probability groups stratified into high and low risk. The dotted lines denote 2-yr and 5-yr RFS/OS and colored annotations show the probability of RFS/OS for each risk group at the respective time points. The shaded areas show the 95% confidence interval (CI) of each curve. A logrank (Mantel-Cox) test was used to assess statistical differences between curves and the logrank method was used to derive hazard ratios. The table below each Kaplan-Meier curve shows the number of events (recurrence events or deaths), total patients (censored+events), and the percentage of events. Bar charts show the pCR rates (see **Fig. 2** legend for descriptions).

*A framework to identify patients for potential de-escalation and escalation strategies*

By stratifying MDX intermediate pCR patients with our MDX-risk signature, two groups emerge with divergent neoadjuvant responses and long-term outcomes: a Favorable Response & Prognosis Profile (FRPP, MDX high pCR + MDX intermediate pCR/MDX-low risk patients) or Adverse Response & Prognosis Profile (ARPP, MDX low pCR + MDX intermediate pCR/MDX-high risk). We explored this Response and Prognosis Profile (RPP) framework in patients with stage II-III TNBC (i.e., those who are eligible for the KN-522 regimen) that are on either the lower or higher spectrum of clinical burden. Patients with lower clinical burden constitute a subset who are more amenable candidates for de-escalation including those with cT1 tumors that have 1-3 positive lymph nodes (cT1/cN1) and cT2-T3 tumors that are node negative. Patients with higher clinical burden may be suitable candidates for escalation or alternative treatment strategies – provided they are predicted to have poor responses to standard of care – and include those with cT1/cN2-3, cT2-T3/N1-3, and cT4/cN0-3 tumors.

Patients with a lower clinical burden and a FRPP had excellent outcomes with only four recurrence events (4/102=3.9%; HR for RFS: 0.16; 95% CI: 0.09-0.29, $p <$ 0.0001; **Supplementary Fig. 8a**) and three deaths (3/106=2.8%; HR for OS: 0.21; 95% CI: 0.10-0.44, $p$ = 0.0044; **Supplementary Fig. 9a**) compared to the ARPP group, which had poorer outcomes with 45 recurrence events (45/190=23.7%) and 27 deaths (27/192=14.1%). Patients with higher clinical burden and a FRPP also had better outcomes relative to those with an ARPP (**Supplementary Fig. 8a, 9a**). Indeed, patients with high clinical burden and an ARPP had higher numbers of recurrence events (62/142=43.7%; HR for RFS: 0.31; 95% CI: 0.20-0.50) and deaths (40/140=28.8%; HR for OS: 0.20; 95% CI: 0.11-0.37). These trends in favorable and adverse long-term outcomes in patients with a FRPP and ARPP, respectively, were largely consistent when further stratified into the CTx+Cb+ICI+ group (**Supplementary Fig. 10a, 11a**) and combined CTx+Cb+ICI− and CTx+Cb−ICI− group (**Supplementary Fig. 12a, 13a**).

*Integration of Response and Prognosis Profiles with TILs*

Given the prognostic utility of TILs in eTNBC, we stratified patients classified as FRPP or ARPP by high and low TILs using a ≥30% cutoff. Patients with a FRPP/High TILs, relative to those with a FRPP/Low TILs, had reduced recurrence rates (6.7% vs 12.2%; **Supplementary Fig. 8b**) although comparable death (3.6% vs 3.9%; **Supplementary Fig. 9b**) rates. Conversely, patients with an ARPP generally had similar RFS/OS outcomes irrespective of TILs, except for lower clinical burden patients, especially those treated with CTx+Cb+ICI− **Supplementary Fig. 8b-13b)**. The synergy of TILs with the RPP framework is also shown in the CTx+Cb+ICI+ (**Supplementary Fig. 10b, 11b**) as well as the CTx+Cb+ICI− and CTx+Cb−ICI− groups (**Supplementary Fig. 12b, 13b**).

*Integration of Response and Prognosis Profiles with neoadjuvant response*

While the MDX RPP captures the biology of baseline chemo-immuno sensitivity and long-term prognoses, this framework could be combined with neoadjuvant response at the time of surgery (pCR or residual disease, RD) to potentially tailor adjuvant treatment regimens. To illustrate this, we stratified patients in the FRPP and ARPP groups by neoadjuvant response or residual disease and assessed changes in RFS and OS. Patients in the FRPP/pCR group, relative to those in the ARPP/pCR group, had fewer recurrence events (8/154=5.19% vs 13/125=10.4%) and deaths (3/158=1.9% vs 7/128=5.5%, **Supplementary Fig. 8c, 9c**). The additional prognostic information that the RPP framework provided in patients with pCR was most evident in patients in the CTx+Cb+ICI− and CTx+Cb−ICI− groups, rather than the CTx+Cb+ICI+ group, which could be due to the longer follow up period in these groups (**Supplementary Fig. 10-13c**). While patients with an ARPP/RD had the worst outcomes, those with a FRPP/RD had more favorable RFS (99/245=40.4% vs 11/55=20.0%) and OS (64/244=26.2% vs 6/57=10.5 %) rates (**Supplementary Fig. 8c, 9c**). The additional prognostic information that the RPP framework provided in patients with RD was again driven by patients in the CTx+Cb+ICI− and CTx+Cb−ICI− groups, especially patients with low clinical burden (**Supplementary Fig. 10-13c**).

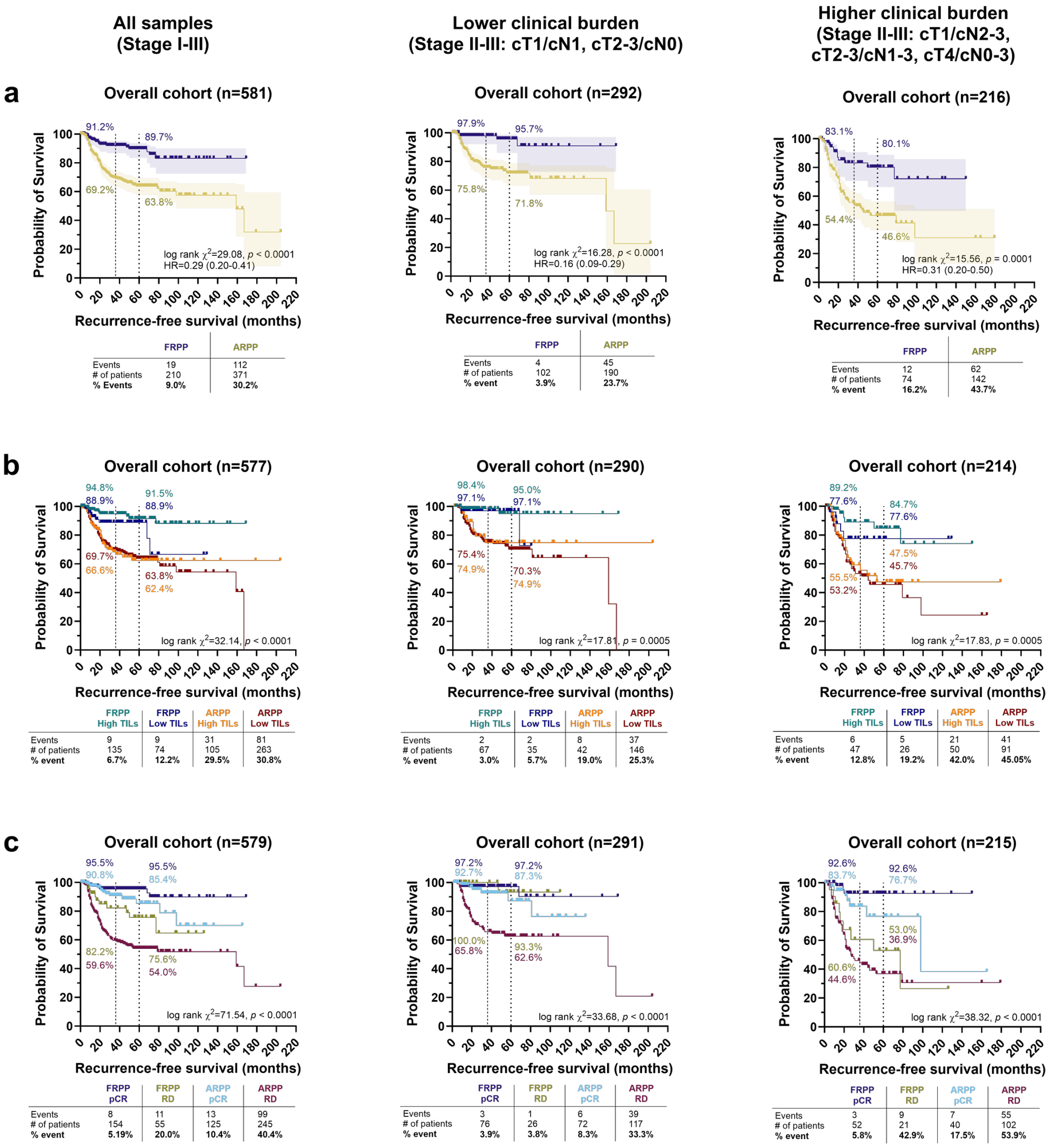


**Supplementary Figure 8.** The MDX Response and Prognosis Profiles integrate with clinical risk level, TILs, and neoadjuvant response to identify patients with disparate recurrence-free survival (RFS). **a-c**) Kaplan-Meier curves show the probability of RFS in **a**) Favorable Response and Prognosis Profile (FRPP) or Adverse Response and Prognosis Profile (ARPP) patients, FRPP or ARPP patients stratified by TILs, and FRPP or ARPP patients stratified by neoadjuvant response (pathological complete response (pCR) or residual disease (RD) at the time of surgery). Left, middle, and right panels **a-c**) show all Stage I-III patients, lower clinical burden Stage II-III patients, and higher clinical burden Stage II-III patients, respectively. The dotted lines denote 3-yr and 5-yr RFS and colored annotations show the probability of RFS for each group at the respective time points. The shaded areas show the 95% confidence interval (CI) of each curve. A logrank (Mantel-Cox) test was used to assess statistical differences between curves and the logrank method was used to derive hazard ratios. The table below each Kaplan-Meier curve shows the number of recurrence events, total patients (censored+recurrence events), and the percentage of recurrence events.

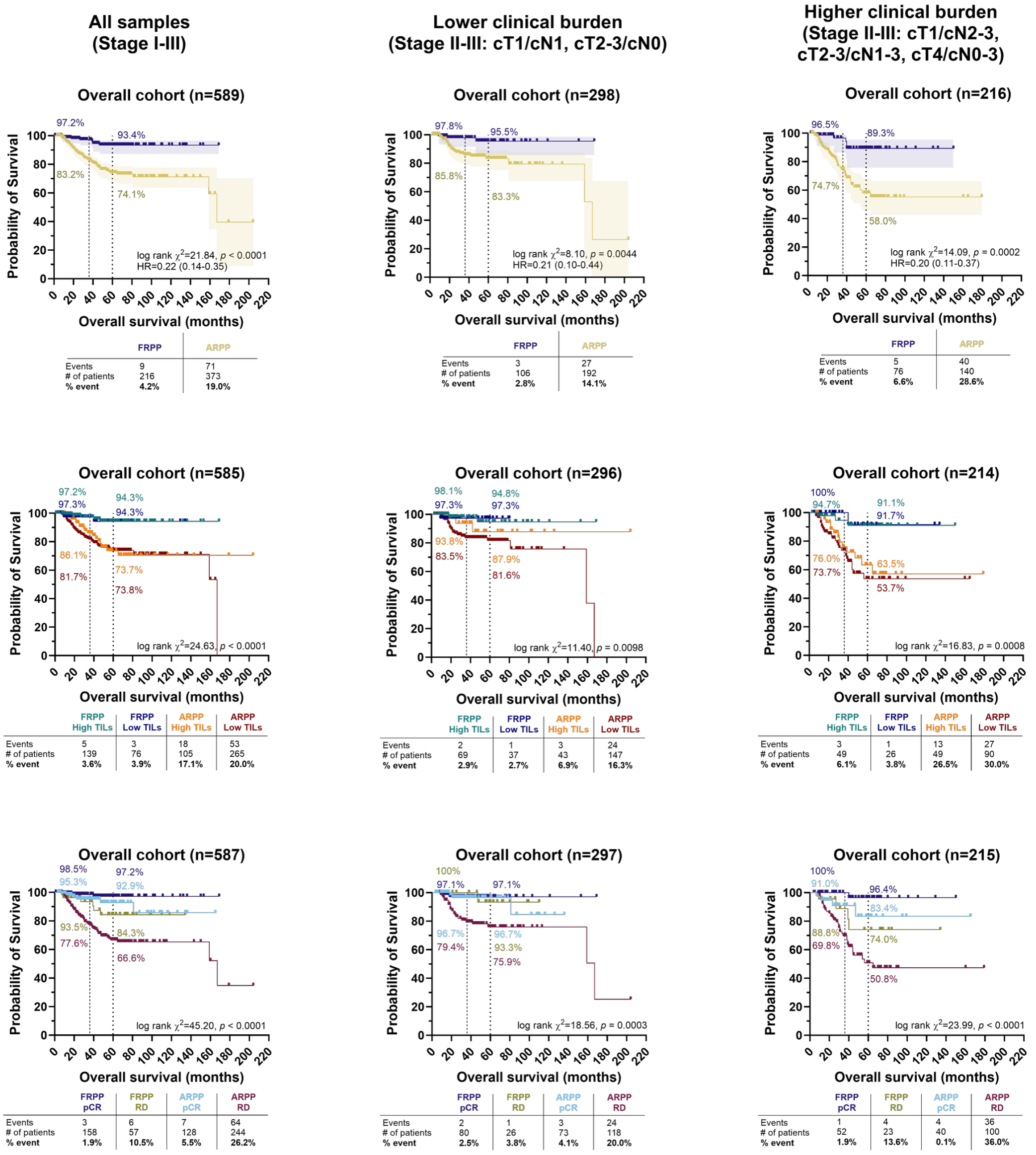


**Supplementary Figure 9.** The MDX Response and Prognosis Profiles integrate with clinical risk level, TILs, and neoadjuvant response to identify patients with disparate overall survival (OS). **a-c**) Kaplan-Meier curves show the probability of OS in **a**) Favorable Response and Prognosis Profile (FRPP) or Adverse Response and Prognosis Profile (ARPP) patients, FRPP or ARPP patients stratified by TILs, and FRPP or ARPP patients stratified by neoadjuvant response (pathological complete response (pCR) or residual disease (RD) at the time of surgery). Left, middle, and right panels **a-c**) show all Stage I-III patients, lower clinical burden Stage II-III patients, and higher clinical burden Stage II-III patients, respectively. The dotted lines denote 2-yr and 5-yr OS and colored annotations show the probability of OS for each group at the respective time points. The shaded areas show the 95% confidence interval (CI) of each curve. A logrank (Mantel-Cox) test was used to assess statistical differences between curves and the logrank method was used to derive hazard ratios. The table below each Kaplan-Meier curve shows the number of events (deaths), total patients (censored+events), and the percentage of events.

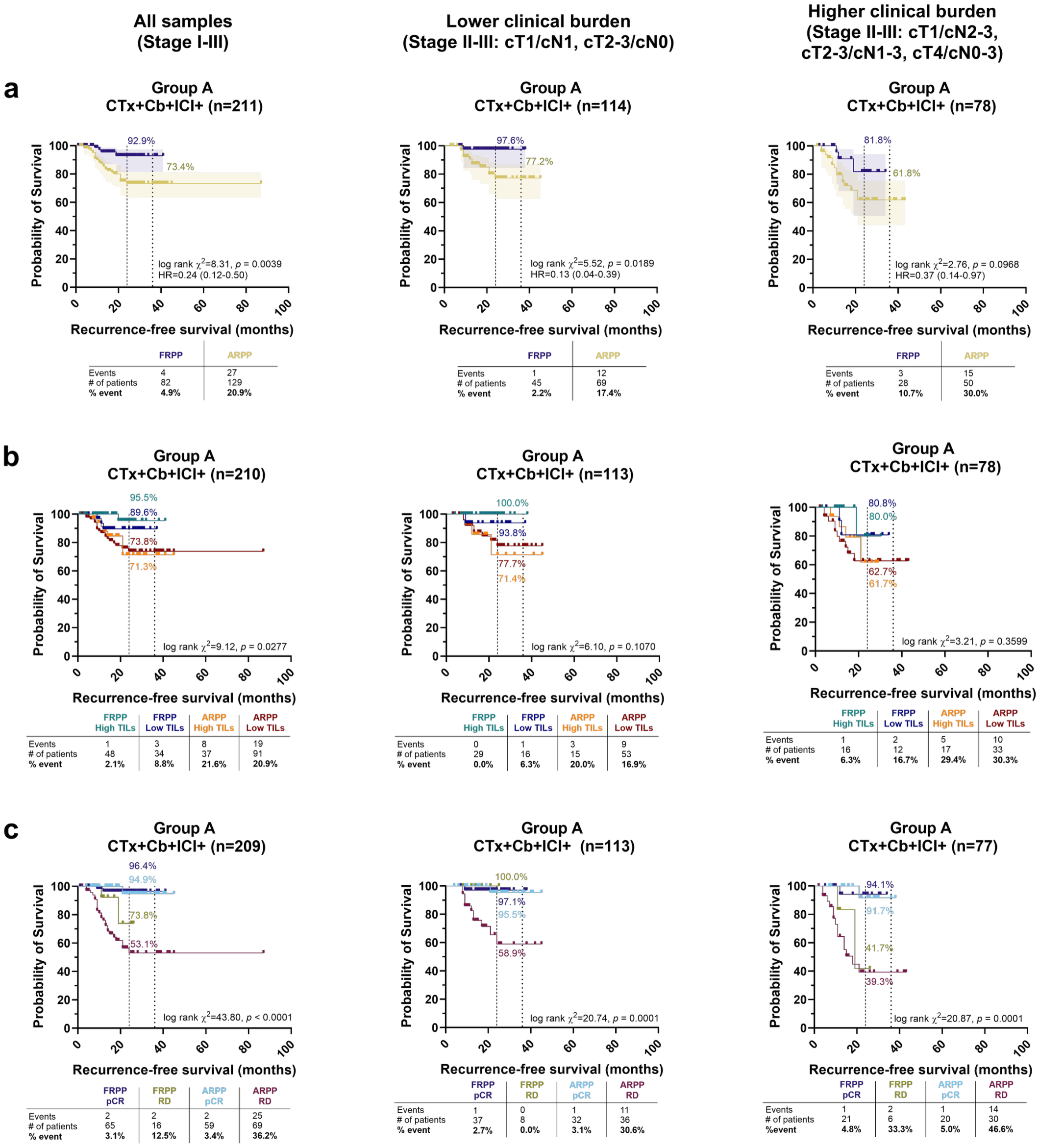


**Supplementary Figure 10.** The MDX Response and Prognosis Profiles integrate with clinical risk level, TILs, and neoadjuvant response to identify immunotherapy-treated patients (group A: CTx+Cb+ICI+) with disparate recurrence-free survival (RFS). **a-c**) Kaplan-Meier curves show the probability of RFS in **a**) Favorable Response and Prognosis Profile (FRPP) or Adverse Response and Prognosis Profile (ARPP) patients, FRPP or ARPP patients stratified by TILs, and FRPP or ARPP patients stratified by neoadjuvant response (pathological complete response (pCR) or residual disease (RD) at the time of surgery). Left, middle, and right panels **a-c**) show all Stage I-III patients, lower clinical burden Stage II-III patients, and higher clinical burden Stage II-III patients, respectively. The dotted lines denote 2-yr and 3-yr RFS and colored annotations show the probability of RFS for each group at the respective time points. The shaded areas show the 95% confidence interval (CI) of each curve. A logrank (Mantel-Cox) test was used to assess statistical differences between curves and the logrank method was used to derive hazard ratios. The table below each Kaplan-Meier curve shows the number of recurrence events, total patients (censored+recurrence events), and the percentage of recurrence events.

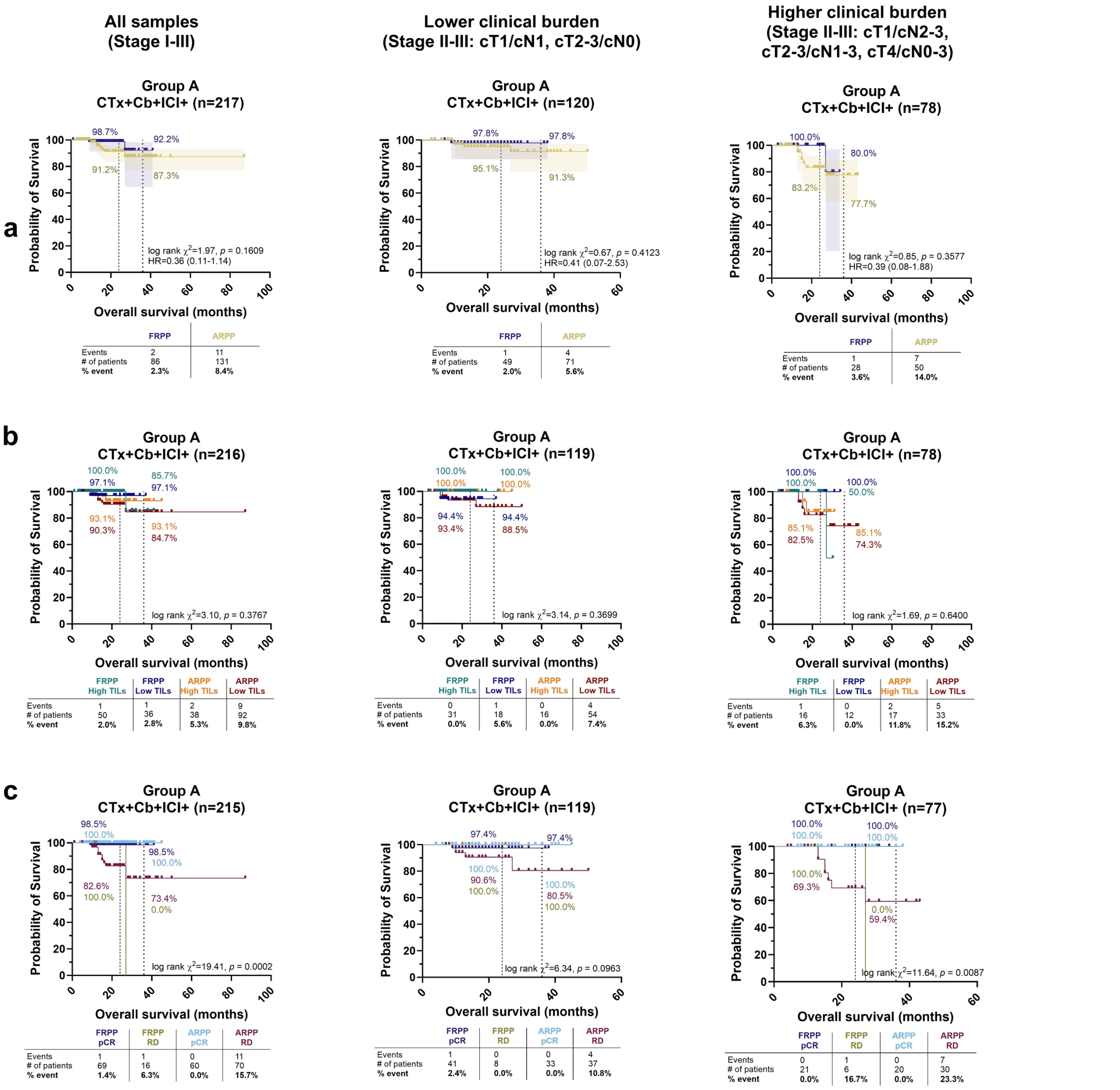


**Supplementary Figure 11.** The MDX Response and Prognosis Profiles integrate with clinical risk level, TILs, and neoadjuvant response to identify immunotherapy-treated patients (group A: CTx+Cb+ICI+) with disparate recurrence-free survival (OS). **a-c**) Kaplan-Meier curves show the probability of OS in **a**) Favorable Response and Prognosis Profile (FRPP) or Adverse Response and Prognosis Profile (ARPP) patients, FRPP or ARPP patients stratified by TILs, and FRPP or ARPP patients stratified by neoadjuvant response (pathological complete response (pCR) or residual disease (RD) at the time of surgery). Left, middle, and right panels **a-c**) show all Stage I-III patients, lower clinical burden Stage II-III patients, and higher clinical burden Stage II-III patients, respectively. The dotted lines denote 2-yr and 3-yr OS and colored annotations show the probability of OS for each group at the respective time points. The shaded areas show the 95% confidence interval (CI) of each curve. A logrank (Mantel-Cox) test was used to assess statistical differences between curves and the logrank method was used to derive hazard ratios. The table below each Kaplan-Meier curve shows the number of events (deaths), total patients (censored+events), and the percentage of events.

| All samples (Stage I-III) | Lower clinical burden (Stage II-III: cT1/cN1, cT2-3/cN0) | Higher clinical burden (Stage II-III: cT1/cN2-3, cT2-3/cN1-3, cT4/cN0-3) |
|---|---|---|

**a**

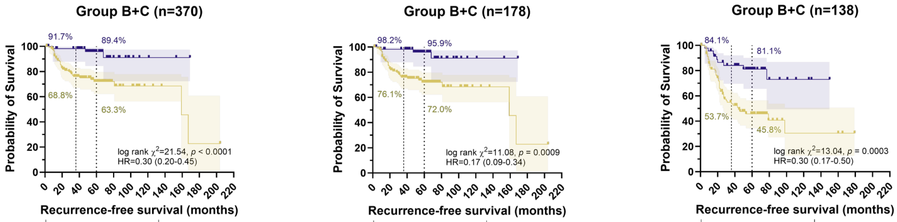


All samples (Stage I-III):

| | | FRPP | ARPP |
|---|---|---|---|
| Groups B+C | Events | 15 | 85 |
| | # of patients | 128 | 242 |
| | % event | **11.7%** | **35.1%** |
| Group B CTx+Cb+ICI– | Events | 3 | 20 |
| | # of patients | 47 | 72 |
| | % event | **6.4%** | **27.8%** |
| Group C CTx+Cb–ICI– | Events | 12 | 65 |
| | # of patients | 81 | 170 |
| | % event | **14.8%** | **38.2%** |

Lower clinical burden:

| | | FRPP | ARPP |
|---|---|---|---|
| Groups B+C | Events | 3 | 33 |
| | # of patients | 57 | 121 |
| | % event | **5.3%** | **27.3%** |
| Group B CTx+Cb+ICI– | Events | 0 | 6 |
| | # of patients | 19 | 38 |
| | % event | **0.0%** | **15.8%** |
| Group C CTx+Cb–ICI– | Events | 3 | 27 |
| | # of patients | 38 | 83 |
| | % event | **7.9%** | **32.5%** |

Higher clinical burden:

| | | FRPP | ARPP |
|---|---|---|---|
| Groups B+C | Events | 9 | 47 |
| | # of patients | 46 | 92 |
| | % event | **19.6%** | **51.1%** |
| Group B CTx+Cb+ICI– | Events | 3 | 13 |
| | # of patients | 17 | 28 |
| | % event | **17.6%** | **46.4%** |
| Group C CTx+Cb–ICI– | Events | 6 | 34 |
| | # of patients | 29 | 64 |
| | % event | **20.7%** | **53.1%** |

**b**

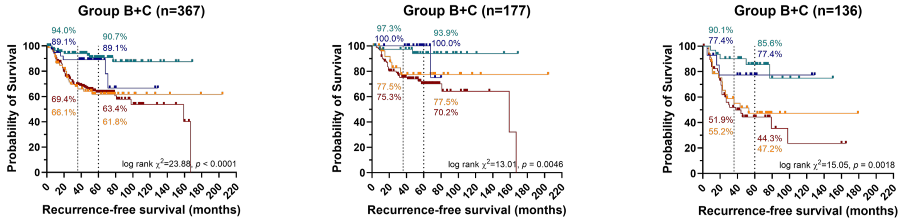


All samples (Stage I-III):

| | | FRPP High TILs | FRPP Low TILs | ARPP High TILs | ARPP Low TILs |
|---|---|---|---|---|---|
| Groups B+C | Events | 8 | 6 | 23 | 62 |
| | # of patients | 87 | 40 | 68 | 172 |
| | % event | **9.2%** | **15.0%** | **33.8%** | **36.0%** |
| Group B CTx+Cb+ICI– | Events | 1 | 1 | 3 | 17 |
| | # of patients | 31 | 15 | 22 | 49 |
| | % event | **3.2%** | **6.7%** | **13.6%** | **34.7%** |
| Group C CTx+Cb–ICI– | Events | 7 | 5 | 20 | 45 |
| | # of patients | 56 | 25 | 46 | 123 |
| | % event | **12.5%** | **20.0%** | **43.5%** | **36.6%** |

Lower clinical burden:

| | | FRPP High TILs | FRPP Low TILs | ARPP High TILs | ARPP Low TILs |
|---|---|---|---|---|---|
| Groups B+C | Events | 2 | 1 | 5 | 28 |
| | # of patients | 38 | 19 | 27 | 93 |
| | % event | **5.3%** | **5.3%** | **18.5%** | **30.1%** |
| Group B CTx+Cb+ICI– | Events | 0 | 0 | 1 | 5 |
| | # of patients | 13 | 6 | 11 | 27 |
| | % event | **0.0%** | **0.0%** | **9.1%** | **18.5%** |
| Group C CTx+Cb–ICI– | Events | 2 | 1 | 4 | 23 |
| | # of patients | 25 | 13 | 16 | 66 |
| | % event | **8.0%** | **7.7%** | **25.0%** | **34.8%** |

Higher clinical burden:

| | | FRPP High TILs | FRPP Low TILs | ARPP High TILs | ARPP Low TILs |
|---|---|---|---|---|---|
| Groups B+C | Events | 5 | 3 | 16 | 31 |
| | # of patients | 31 | 14 | 33 | 58 |
| | % event | **16.1%** | **21.4%** | **48.5%** | **53.4%** |
| Group B CTx+Cb+ICI– | Events | 1 | 1 | 2 | 11 |
| | # of patients | 10 | 6 | 9 | 18 |
| | % event | **10.0%** | **16.7%** | **22.2%** | **61.1%** |
| Group C CTx+Cb–ICI– | Events | 4 | 2 | 14 | 20 |
| | # of patients | 21 | 8 | 24 | 40 |
| | % event | **19.0%** | **25.0%** | **58.3%** | **50.0%** |

**c**

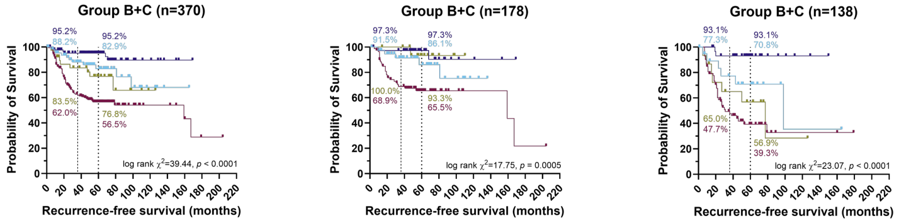


All samples (Stage I-III):

| | | FRPP pCR | FRPP RD | ARPP pCR | ARPP RD |
|---|---|---|---|---|---|
| Groups B+C | Events | 6 | 9 | 11 | 74 |
| | # of patients | 89 | 39 | 66 | 176 |
| | % event | **6.7%** | **23.1%** | **16.7%** | **42.0%** |
| Group B CTx+Cb+ICI– | Events | 1 | 2 | 3 | 17 |
| | # of patients | 36 | 11 | 25 | 47 |
| | % event | **2.8%** | **18.2%** | **12.0%** | **36.2%** |
| Group C CTx+Cb–ICI– | Events | 5 | 7 | 8 | 57 |
| | # of patients | 53 | 28 | 41 | 129 |
| | % event | **9.4%** | **25.0%** | **19.5%** | **44.2%** |

Lower clinical burden:

| | | FRPP pCR | FRPP RD | ARPP pCR | ARPP RD |
|---|---|---|---|---|---|
| Groups B+C | Events | 2 | 1 | 5 | 28 |
| | # of patients | 39 | 18 | 40 | 81 |
| | % event | **5.1%** | **5.5%** | **12.5%** | **34.6%** |
| Group B CTx+Cb+ICI– | Events | 0 | 0 | 2 | 4 |
| | # of patients | 15 | 4 | 17 | 21 |
| | % event | **0.0%** | **0.0%** | **11.8%** | **19.0%** |
| Group C CTx+Cb–ICI– | Events | 2 | 1 | 3 | 24 |
| | # of patients | 24 | 14 | 23 | 60 |
| | % event | **8.3%** | **7.1%** | **13.0%** | **40.0%** |

Higher clinical burden:

| | | FRPP pCR | FRPP RD | ARPP pCR | ARPP RD |
|---|---|---|---|---|---|
| Groups B+C | Events | 2 | 7 | 6 | 41 |
| | # of patients | 31 | 15 | 20 | 72 |
| | % event | **6.5%** | **46.7%** | **30.0%** | **56.9%** |
| Group B CTx+Cb+ICI– | Events | 1 | 2 | 1 | 12 |
| | # of patients | 13 | 4 | 7 | 21 |
| | % event | **7.7%** | **50.0%** | **14.3%** | **57.1%** |
| Group C CTx+Cb–ICI– | Events | 1 | 5 | 5 | 29 |
| | # of patients | 18 | 11 | 13 | 51 |
| | % event | **5.6%** | **45.5%** | **38.5%** | **56.9%** |

**Supplementary Figure 12.** The MDX Response and Prognosis Profiles integrate with clinical risk level, TILs, and neoadjuvant response to identify chemotherapy-treated patients (groups B+C: CTx+Cb+ICI− and CTx+Cb−ICI−) with disparate recurrence-free survival (RFS). **a-c**) Kaplan-Meier curves show the probability of RFS in **a**) Favorable Response and Prognosis Profile (FRPP) or Adverse Response and Prognosis Profile (ARPP) patients, FRPP or ARPP patients stratified by TILs, and FRPP or ARPP patients stratified by neoadjuvant response (pathological complete response (pCR) or residual disease (RD) at the time of surgery). Left, middle, and right panels **a-c**) show all Stage I-III patients, lower clinical burden Stage II-III patients, and higher clinical burden Stage II-III patients, respectively. The dotted lines denote 3-yr and 5-yr RFS and colored annotations show the probability of RFS for each group at the respective time points. The shaded areas show the 95% confidence interval (CI) of each curve. A logrank (Mantel-Cox) test was used to assess statistical differences between curves and the logrank method was used to derive hazard ratios. The table below each Kaplan-Meier curve shows the number of recurrence events, total patients (censored+recurrence events), and the percentage of recurrence events.

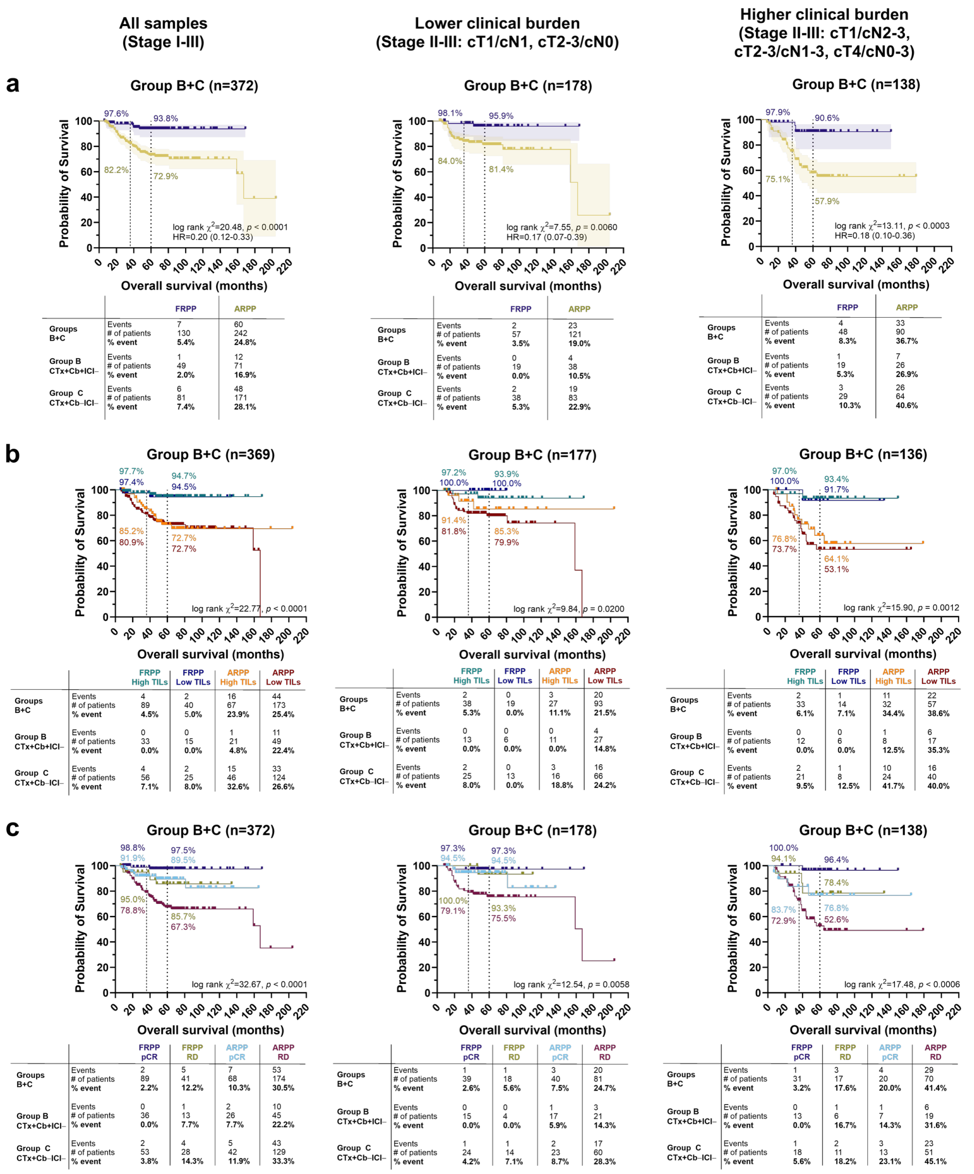


**a) All samples**

| Groups | | FRPP | ARPP |
|---|---|---|---|
| Groups B+C | Events | 7 | 60 |
| | # of patients | 130 | 242 |
| | % event | 5.4% | 24.8% |
| Group B CTx+Cb+ICI− | Events | 1 | 12 |
| | # of patients | 49 | 71 |
| | % event | 2.0% | 16.9% |
| Group C CTx+Cb−ICI− | Events | 6 | 48 |
| | # of patients | 81 | 171 |
| | % event | 7.4% | 28.1% |

**a) Lower clinical burden**

| Groups | | FRPP | ARPP |
|---|---|---|---|
| Groups B+C | Events | 2 | 23 |
| | # of patients | 57 | 121 |
| | % event | 3.5% | 19.0% |
| Group B CTx+Cb+ICI− | Events | 0 | 4 |
| | # of patients | 19 | 38 |
| | % event | 0.0% | 10.5% |
| Group C CTx+Cb−ICI− | Events | 2 | 19 |
| | # of patients | 38 | 83 |
| | % event | 5.3% | 22.9% |

**a) Higher clinical burden**

| Groups | | FRPP | ARPP |
|---|---|---|---|
| Groups B+C | Events | 4 | 33 |
| | # of patients | 48 | 90 |
| | % event | 8.3% | 36.7% |
| Group B CTx+Cb+ICI− | Events | 1 | 7 |
| | # of patients | 19 | 26 |
| | % event | 5.3% | 26.9% |
| Group C CTx+Cb−ICI− | Events | 3 | 26 |
| | # of patients | 29 | 64 |
| | % event | 10.3% | 40.6% |

**b) All samples**

| Groups | | FRPP High TILs | FRPP Low TILs | ARPP High TILs | ARPP Low TILs |
|---|---|---|---|---|---|
| Groups B+C | Events | 4 | 2 | 16 | 44 |
| | # of patients | 89 | 40 | 67 | 173 |
| | % event | 4.5% | 5.0% | 23.9% | 25.4% |
| Group B CTx+Cb+ICI− | Events | 0 | 0 | 1 | 11 |
| | # of patients | 33 | 15 | 21 | 49 |
| | % event | 0.0% | 0.0% | 4.8% | 22.4% |
| Group C CTx+Cb−ICI− | Events | 4 | 2 | 15 | 33 |
| | # of patients | 56 | 25 | 46 | 124 |
| | % event | 7.1% | 8.0% | 32.6% | 26.6% |

**b) Lower clinical burden**

| Groups | | FRPP High TILs | FRPP Low TILs | ARPP High TILs | ARPP Low TILs |
|---|---|---|---|---|---|
| Groups B+C | Events | 2 | 0 | 3 | 20 |
| | # of patients | 38 | 19 | 27 | 93 |
| | % event | 5.3% | 0.0% | 11.1% | 21.5% |
| Group B CTx+Cb+ICI− | Events | 0 | 0 | 0 | 4 |
| | # of patients | 13 | 6 | 11 | 27 |
| | % event | 0.0% | 0.0% | 0.0% | 14.8% |
| Group C CTx+Cb−ICI− | Events | 2 | 0 | 3 | 16 |
| | # of patients | 25 | 13 | 16 | 66 |
| | % event | 8.0% | 0.0% | 18.8% | 24.2% |

**b) Higher clinical burden**

| Groups | | FRPP High TILs | FRPP Low TILs | ARPP High TILs | ARPP Low TILs |
|---|---|---|---|---|---|
| Groups B+C | Events | 2 | 1 | 11 | 22 |
| | # of patients | 33 | 14 | 32 | 57 |
| | % event | 6.1% | 7.1% | 34.4% | 38.6% |
| Group B CTx+Cb+ICI− | Events | 0 | 0 | 1 | 6 |
| | # of patients | 12 | 6 | 8 | 17 |
| | % event | 0.0% | 0.0% | 12.5% | 35.3% |
| Group C CTx+Cb−ICI− | Events | 2 | 1 | 10 | 16 |
| | # of patients | 21 | 8 | 24 | 40 |
| | % event | 9.5% | 12.5% | 41.7% | 40.0% |

**c) All samples**

| Groups | | FRPP pCR | FRPP RD | ARPP pCR | ARPP RD |
|---|---|---|---|---|---|
| Groups B+C | Events | 2 | 5 | 7 | 53 |
| | # of patients | 89 | 41 | 68 | 174 |
| | % event | 2.2% | 12.2% | 10.3% | 30.5% |
| Group B CTx+Cb+ICI− | Events | 0 | 1 | 2 | 10 |
| | # of patients | 36 | 13 | 26 | 45 |
| | % event | 0.0% | 7.7% | 7.7% | 22.2% |
| Group C CTx+Cb−ICI− | Events | 2 | 4 | 5 | 43 |
| | # of patients | 53 | 28 | 42 | 129 |
| | % event | 3.8% | 14.3% | 11.9% | 33.3% |

**c) Lower clinical burden**

| Groups | | FRPP pCR | FRPP RD | ARPP pCR | ARPP RD |
|---|---|---|---|---|---|
| Groups B+C | Events | 1 | 1 | 3 | 20 |
| | # of patients | 39 | 18 | 40 | 81 |
| | % event | 2.6% | 5.6% | 7.5% | 24.7% |
| Group B CTx+Cb+ICI− | Events | 0 | 0 | 1 | 3 |
| | # of patients | 15 | 4 | 17 | 21 |
| | % event | 0.0% | 0.0% | 5.9% | 14.3% |
| Group C CTx+Cb−ICI− | Events | 1 | 1 | 2 | 17 |
| | # of patients | 24 | 14 | 23 | 60 |
| | % event | 4.2% | 7.1% | 8.7% | 28.3% |

**c) Higher clinical burden**

| Groups | | FRPP pCR | FRPP RD | ARPP pCR | ARPP RD |
|---|---|---|---|---|---|
| Groups B+C | Events | 1 | 3 | 4 | 29 |
| | # of patients | 31 | 17 | 20 | 70 |
| | % event | 3.2% | 17.6% | 20.0% | 41.4% |
| Group B CTx+Cb+ICI− | Events | 0 | 1 | 1 | 6 |
| | # of patients | 13 | 6 | 7 | 19 |
| | % event | 0.0% | 16.7% | 14.3% | 31.6% |
| Group C CTx+Cb−ICI− | Events | 1 | 2 | 3 | 23 |
| | # of patients | 18 | 11 | 13 | 51 |
| | % event | 5.6% | 18.2% | 23.1% | 45.1% |

**Supplementary Figure 13.** The MDX Response and Prognosis Profiles integrate with clinical risk level, TILs, and neoadjuvant response to identify chemotherapy-treated patients (groups B+C: CTx+Cb+ICI− and CTx+Cb−ICI−) with disparate recurrence-free survival (OS). **a-c**) Kaplan-Meier curves show the probability of OS in **a**) Favorable Response and Prognosis Profile (FRPP) or Adverse Response and Prognosis Profile (ARPP) patients, FRPP or ARPP patients stratified by TILs, and FRPP or ARPP patients stratified by neoadjuvant response (pathological complete response (pCR) or residual disease (RD) at the time of surgery). Left, middle, and right panels **a-c**) show all Stage I-III patients, lower clinical burden Stage II-III patients, and higher clinical burden Stage II-III patients, respectively. The dotted lines denote 3-yr and 5-yr OS and colored annotations show the probability of OS for each group at the respective time points. The shaded areas show the 95% confidence interval (CI) of each curve. A logrank (Mantel-Cox) test was used to assess statistical differences between curves and the logrank method was used to derive hazard ratios. The table below each Kaplan-Meier curve shows the number of events (deaths), total patients (censored+events), and the percentage of events.
